# Bacillus Calmette-Guérin vaccine to reduce COVID-19 infections and hospitalisations in healthcare workers – a living systematic review and prospective ALL-IN meta-analysis of individual participant data from randomised controlled trials

**DOI:** 10.1101/2022.12.15.22283474

**Authors:** J.A. (Judith) ter Schure, Alexander Ly, Lisa Belin, Christine S. Benn, Marc J.M. Bonten, Jeffrey D. Cirillo, Johanna A.A. Damen, Inês Fronteira, Kelly D. Hendriks, Ana Paula Junqueira-Kipnis, André Kipnis, Odile Launay, Jose Euberto Mendez-Reyes, Judit Moldvay, Mihai G. Netea, Sebastian Nielsen, Caryn M. Upton, Gerben van den Hoogen, Jesper M. Weehuizen, Peter D. Grünwald, C.H. (Henri) van Werkhoven

**Affiliations:** Amsterdam UMC/CWI, Amsterdam, the Netherlands; University of Amsterdam/CWI, Amsterdam, the Netherlands; Sorbonne Université, INSERM, Institut Pierre Louis d’Epidémiologie et de Santé Publique, AP-HP, Hôpital Pitié Salpêtrière, Département de Santé Publique, Unité de Recherche Cli-nique PSL-CFX, CIC-1901, F75013, Paris, France; Bandim Health Project, Open Patient Data Explorative Network, Department of Clinical Research and Danish Institute for Advanced Study, University of Southern Denmark, Denmark; Julius Center for Health Sciences and Primary Care, University Medical Center Utrecht, Utrecht University, Utrecht, the Netherlands; Center for Airborne Pathogen Research and Imaging, Texas A&M School of Medicine, Bryan, TX 77807; Cochrane Netherlands, Julius Center for Health Sciences and Primary Care, University Medical Center Utrecht, Utrecht University, Utrecht, the Netherlands; Global Health and Tropical Medicine, Institute of Hygiene and Tropical Medicine, Universidade NOVA de Lisboa; Federal University of Goiás, Institute of Tropical Medicine and Public Health, Goiânia, Brazil; AP-HP; Global and Immigrant Health, Baylor College of Medicine, Houston, TX; National Korányi Institute of Pulmonology, Budapest, Hungary; Department of Internal Medicine and Radboud Center for Infectious Diseases, Radboud University Medical Center, Nijmegen, the Netherlands; Bandim Health Project, Open Patient Data Explorative Network, Department of Clinical Research, University of Southern Denmark, Denmark; TASK, Parow, Cape Town, South Africa; Department of Internal Medicine and Infectious Diseases, University Medical Center Utrecht, Utrecht University, Utrecht, the Netherlands; CWI, Amsterdam, the Netherlands

## Abstract

**BACKGROUND:** The objective is to determine the impact of the Bacillus Calmette*-*Guérin (BCG) vaccine compared to placebo or no vaccine on COVID-19 infections and hospitalisations in healthcare workers. We are using a living and prospective approach to Individual-Participant-Data (IPD) meta-analysis of ongoing studies based on the Anytime Live and Leading Interim (ALL-IN) meta-analysis statistical methodology.

**METHODS:** Planned and ongoing randomised controlled trials were identified from trial registries and by snowballing (final elicitation: Oct 3 2022). The methodology was specified prospectively – with no trial results available – for trial inclusion as well as statistical analysis. Inclusion decisions were made collaboratively based on a risk-of-bias assessment by an external protocol review committee (Cochrane risk-of-bias tool adjusted for use on protocols), expected homogeneity in treatment effect, and agreement with the predetermined event definitions. The co-primary endpoints were incidence of COVID-19 infection and COVID-19-related hospital admission. Accumulating IPD from included trials was analysed sequentially using the exact *e*-value logrank test (at level α = 0.5% for infections and level α = 4.5% for hospitalisations) and anytime-valid 95%-confidence intervals (CIs) for the hazard ratio (HR) for a predetermined fixed-effects approach to meta-analysis (no measures of statistical heterogeneity). Infections were included if demonstrated by PCR tests, antigen tests or suggestive lung CTs. Participants were censored at date of first COVID-19-specific vaccination and two-stage analyses were performed in calendar time, with a stratification factor per trial.

**RESULTS:** Six trials were included in the primary analysis with 4 433 participants in total. The *e*-values showed no evidence of a favourable effect of minimal clinically relevance (HR < 0.8) in comparison to the null (HR = 1) for COVID-19 infections, nor for COVID-19 hospitalisations (HR < 0.7 vs HR = 1). COVID-19 infection was observed in 251 participants receiving BCG and 244 participants not receiving BCG, HR 1.02 (anytime-valid 95%-CI 0.78-1.35). COVID-19 hospitalisations were observed in 13 participants receiving BCG and 7 not receiving BCG, resulting in an uninformative estimate (HR 1.88; anytime-valid 95%-CI 0.26-13.40).

**DISCUSSION:** It is highly unlikely that BCG has a clinically relevant effect on COVID-19 infections in healthcare workers. With only limited observations, no conclusion could be drawn for COVID-19 related hospitalisation. Due to the nature of ALL-IN meta-analysis, emerging data from new trials can be included without violating type*-*I error rates or interval coverage. We intend to keep this meta-analysis alive and up-to-date, as more trials report. For COVID-19 related hospitalisations, we do not expect enough future observations for a meaningful analysis. For BCG-mediated protection against COVID-19 infections, on the other hand, more observations could lead to a more precise estimate that concludes the meta-analysis for futility, meaning that the current interval excludes the HR of 0.8 predetermined as effect size of minimal clinical relevance.

**OTHER:** No external funding. Preregistered at PROSPERO: CRD42021213069.

## Introduction

With the emergence of Severe Acute Respiratory Syndrome Coronavirus 2 (SARS-CoV-2) in 2019, leading to the Coronavirus disease 2019 (COVID-19) pandemic in early 2020, a global search for effective prevention and treatment modalities was initiated. Earlier epidemiological studies and clinical trials have suggested that Bacillus Calmette*-*Guérin (BCG) vaccine induces heterologous protection against respiratory tract infections in both young children and adults (Wardhana, E.A., Sultana, Mandang, & Jim, 2011; Aaby, et al., 2011; de Castro, Pardo-Seco, & Martinón-Torres, 2015; Nemes, Geldenhuys, Rozot, & others, 2018; Giamarellos-Bourboulis, Tsilika, S, & al., 2020; Singh, Netea, & Bishai, 2021). The mechanisms of protection have been suggested to involve induction of heterologous T-cell responses (Benn, Netea, Selin, & Aaby, 2013) and innate immune reprogramming (also termed *trained immunity* (Netea, et al., 2020)). Supported by ecological studies, suggesting lower COVID-19 incidences in countries with active national BCG vaccination policy (O’Connor, Teh, Kamat, & Lawrentschuk, 2020), researchers across the world initiated randomised controlled trials with BCG as a non-specific protective approach against COVID-19. A substantial number of these trials targeted healthcare workers because of their high exposure rates and fear of a healthcare crisis due to high absenteeism rates. Many of these trials were very similar in their design, especially when they were inspired by one of the first trials (Ten Doesschate, et al., 2022) that had shared their protocol.

Having multiple independent trials in parallel increases the long run precision of effect estimates and improves generalisability of results. However, the situation poses a risk of false*-*positive or too-late findings. On the one hand, with over 20 ongoing trials and many of these performing interim analyses, the probability that at least one such trial would find superiority early compared to available results in other trials, under the hypothesis of no effect, is much larger than the typically accepted type*-*I error rate of 5%. Such a scenario might have direct implications for ongoing trials such as a decision to stop follow-up and vaccinate the control group without knowledge of the totality of accumulated evidence. On the other hand, the power of each trial was uncertain due to the pandemic situation with large fluctuations in COVID-19 incidence. Each trial separately could be underpowered, especially for identifying effects on severe disease, and a protective effect might only be observed after trial data were combined. The pandemic situation at the time urged for a collaborative approach with a statistical method that enables continuous synthesis of the data from all ongoing trials.

It can take months or years to collect and analyse the data of all trials and perform an individual-participant-data (IPD) meta-analysis (Tierney, Riley, Smith, Clarke, & Stewart, 2021). By simplifying the structure of the data collection and inviting all ongoing trials, the current study aimed to collect accumulating data and perform an Anytime Live and Leading INterim (ALL-IN) meta-analysis. The results presented here are based on the limited data that was accumulated on an ongoing basis. The analysis is rich in its representation of the history of the evidence and timing of events, yet it does not contain all the (subgroup/covariate-adjusted) analyses that would be possible by combining all full trial datasets.

Our collaboration was named ALL-IN-META-BCG-CORONA. *Figure 1* illustrates what the approach was designed to achieve. In this hypothetical case, not only could the world benefit much earlier from the knowledge of an effective existing vaccine in a pandemic, the initiation of hypothetical trial 7 could have been advised against, reducing possible research waste (Chalmers & Glasziou, 2009; Glasziou, Sanders, & Hoffmann, 2020).

**Figure 1.**
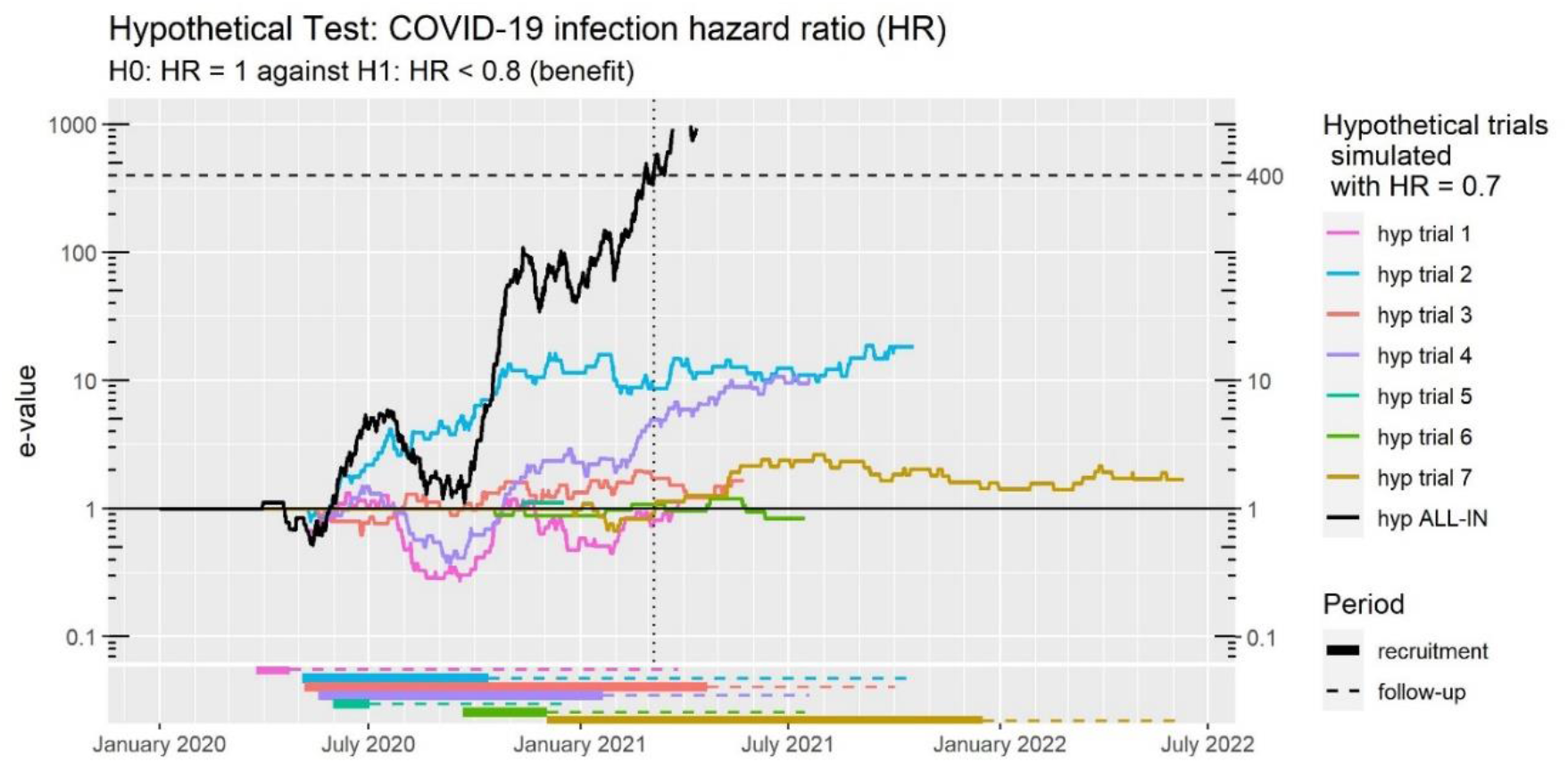
E-values for seven hypothetical trials. These are e-values that test hazard ratio 1 versus 0.8 and are explained in the **Methods** section below, that also addresses the accompanying threshold of 400 in more detail in section **E-value analysis design: effect sizes of minimal clinical relevance and thresholds**. Note that the y-axis is logarithmic.

## Methods

### Aims of ALL-IN-META-BCG-CORONA

The aim is to keep track of all ongoing trials by calculating for each calendar date and each trial a notion of evidence – the *e*-value. As illustrated by *Figure 1* for the analysis of events of COVID-19 infection, the sequence of *e*-values goes up for each observed event in the control group (indicating possible protection in the BCG group) and goes down for each observed event in the BCG group (indicating the opposite). The e-value would therefore accumulate evidence indicating either a protective effect of BCG (increasing *e*-value), or no or even a harmful effect (decreasing *e*-value). Each trial contributes *e*-values from the first observed event until the last, and these *e-*values are combined in a meta-analysis *e-*value by multiplication, as described in the paper introducing the ALL-IN approach (Ter Schure & Grünwald, 2022) and the more general literature on combining *e*-values (Vovk & Wang, 2021).

Since the meta-analysis *e-*value is based on more events than any individual trial alone, it can reach higher levels of evidence earlier. *Figure 1* shows that this hypothetical case with seven trials where the evidence reaches a level of 400 in March 2021. At that time, most of these hypothetical trials still have participants in follow-up with two actively recruiting (hyp trial 3 and 7). The *e-*values allow for the meta-analysis to be the leading source of information for any decisions to start, stop or expand trials, e.g. advising against the initiation of hypothetical trial 7.

### E-values and anytime-valid confidence intervals

ALL-IN-META-BCG-CORONA is based on *e*-values for hypothesis testing (*Figure 1*). Their counterpart for estimation is the anytime-valid confidence interval. These methods have recently seen further development – since early work in the ‘40s and ‘60s-’80s (Stein & Wald, 1947; Darling & Robbins, 1967) – and attracted considerable attention in the statistics literature (Wasserman, Ramdas, & Balakrishnan, 2020; Vovk & Wang, 2021; Shafer, 2021; Howard, Ramdas, McAuliffe, & Sekhon, 2021; Henzi & Ziegel, 2022; Grünwald, De Heide, & Koolen, 2022). They distinguish themselves from methods based on *p*-vales and conventional confidence intervals by the fact that they can be continuously updated without losing statistical validity.

While a small *p*-value is better, a large *e*-value is better. Broadly speaking, *e=7* indicates seven times more evidence for the hypothesis that BCG has a favourable effect compared to the null. We can set a threshold for a predetermined amount of evidence to keep an eye out for, like *e* > 400 in *Figure 1*. A threshold of 400 controls type-I error at a level *α*= 1/400 = 0.25%, and can be used to inform decisions about the data collection in ongoing and new trials while retaining that type-I error control (Ter Schure & Grünwald, 2022). For more details, see the Appendix section ***Detailed methods***.

### IPD, living, interim, bottom-up, prospective and collaborative meta-analysis

*E*-values and anytime-valid confidence intervals can be used in many different meta-analysis settings, such as those based on IPD or aggregate data, final analysis or living systematic review, complete trials or interim, top-down (like a multi-centre trial) or bottom-up, retrospective or prospective, external or collaborative (Ter Schure & Grünwald, 2022). ALL-IN-META-BCG-CORONA is (a) IPD, (b) living, (c) interim, (d) bottom-up (but also slightly top-down, see below), (e) prospective and (f) collaborative.

#### (a) Individual Participant Data (IPD); reducing data problems

The data was shared in file formats with a row of data per participant. With full information on the date of randomisation, events and follow-up, the evolution of evidence over time could be retrospectively processed, based on each new data upload. Only information for the time-to-event analysis in the two primary outcomes (COVID-19 infections and hospitalisations) was collected, along with information on the stratifying factor, limiting the amount of work in ongoing data cleaning. The IPD approach encourages close collaboration between the trial data-uploaders and meta-analysis statisticians. This improves problem solving in data cleaning and optimises data quality. The IPD approach also limits the risk of falsified or fabricated data entering the meta-analysis because more than one person inspects all the data.

#### (b) Living and on (c) Interim data

New trials were contacted as soon as we became aware of their existence. Crucially, we did not only analyse completed trials, but included interim datasets to aim for a live account of the evidence while the trials were ongoing.

#### (d) Bottom-up (but also slightly top-down); enabling homogeneity

Each trial could specify their own stratification factor (‘hospital’ see ***Data extraction***) in the data uploaded, either following their randomisation strategy for multi-centre trials that randomised stratified by hospital of occupation, or based on other available information that determined common risk of COVID-19. In this sense, part of the data analysis decisions are bottom-up. Also, in contrast to methods based on group-sequential and alpha-spending approaches (Simmonds, et al., 2017), ALL ALL-IN meta-analysis does not require a predetermined stopping boundary. There is a threshold, but rather than being a top-down decision rule, this merely serves for optimal timing to consider stopping, while the analysis stays valid if data collection continues. The specific *e*-values used, however, do require a predetermined effect size of minimal clinical relevance (Grünwald, De Heide, & Koolen, 2022), which was decided top-down by the meta-analysis steering committee (see further information in section ***E-value analysis design: effect sizes of minimal clinical relevance*** and the Appendix section ***Detailed methods***).

#### (e) Prospective; minimizing bias

All decisions on effect size of minimal clinical relevance (Van Werkhoven, et al., 2021), event definitions and trial inclusion were made before results were known, as prescribed for a prospective meta-analysis (Seidler, et al., 2019). This reduced risks of publication bias and other meta-analysis “significance-chasing biases” (Ioannidis, 2010). In agreement with this recommendation by the Cochrane PMA Methods Group (Seidler, et al., 2019), we constructed a central steering and data analysis committee, and worked in collaboration with representatives (the PI and a data-uploader) from each individual study.

#### (f) Collaborative; using a dashboard

All trials were represented by a team member in an Advisory committee and members were involved in the meta-analysis while their trials were ongoing, and in some cases, while their trials were still in preparation. This allowed for detailed information sharing about the trials to inform decision making. Moreover, it also facilitated direct information flow from the meta-analysis to the trial members becoming the leading source of information on decisions to start, stop or expand trials. A dashboard, shown in demo-mode in Figure 2, was used to communicate the *e*-values throughout the course of the participating trials. Dashboard access permissions were regulated based on the stage of blinding each trial was in, with initial permissions only granted to data uploaders unblinded to their own trial results (with other trials not visible), and later all trials were made visible to all data-uploaders and participating investigators. The launch of the dashboard occurred once three trials were ready to be included, such that initially no individual trial contributions could be retrofitted from the meta-analysis *e-*values and a single other trial’s *e-*values.

**Figure 2.**
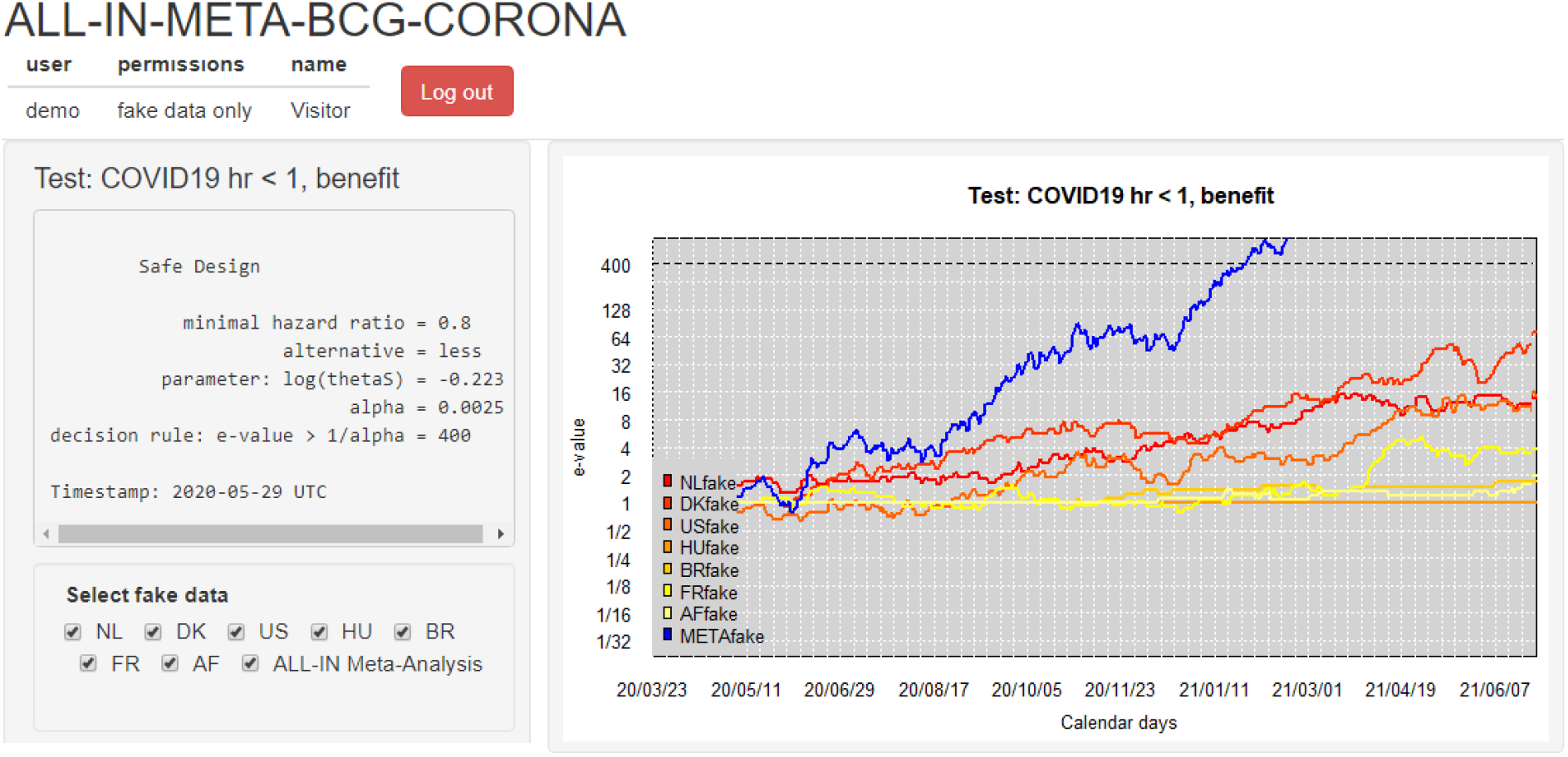
Dashboard used to communicate meta-analysis results to all data-uploaders with a login. The dashboard is in demo mode, showing synthetic (‘fake’) data until June 2021. The option to (de)select trials is for plotting purposes of individual trial e-values; all trials in the dashboard stay included in the meta e-value, following the decision on trial inclusion. The demo dashboard was openly available to easily explain the project to outsiders, using user name ‘demo’, password ‘show’ (Ter Schure J., ALL-IN-META-BCG-CORONA dashboard, 2020). Note that the y-axis is on the log scale.

### Structure of the collaboration

Our collaboration consists of a Steering committee (PDG, MGN, MJMB), a representative of Cochrane Netherlands (JAAD) with two collaborators (KDH and JMW), an Advisory committee including a principal investigator and data uploader representing each trial, and an Operational team (JAS, AL, CHW), with the first two acting as meta-analysis statisticians and the last as meta principal investigator.

The Steering committee stayed blinded to the interim results until all trials were concluded and (1) decided on the primary outcome measures, effect size of minimal clinical relevance, thresholds, and event definitions, (2) decided on trial inclusion, advised by the Advisory committee and Cochrane Netherlands, and (3) decided when to make the meta-analysis results public in the dashboard and/or in a scientific publication by consulting the Advisory committee.

Cochrane Netherlands performed an external risk-of-bias assessment based on the protocols and feedback from the participating trials.

The Advisory committee (1) provided Cochrane Netherlands with detailed protocol information to perform a risk-of-bias assessment, (2) advised on trial inclusion criteria, and (3) advised on when to make the meta-analysis results public.

The Operational team (1) identified the trials, (2) coordinated data collection, (3) analysed data and updated the dashboard, (4) wrote news updates, and (5) prepared the publication.

All documents detailing this approach can be found in the supplementary material available at Research Equals (ALL-IN-META-BCG-CORONA Replication Package). These include the Statistical Analysis Plan, a webinar and tutorials explaining the statistical approach and analysis code, newsletters with updates throughout the pandemic, risk-of-bias assessments, summary data and links to data publications.

### Identification of trials

To identify trials for inclusion in the meta-analysis, ClinicalTrials.gov was searched for the terms “BCG AND (COVID OR corona OR SARS-CoV-2)”. We also screened a database constructed for the Kaggle hackathon on BCG and COVID-19 clinical trials (Kaggle, 2020). Finally, we used snowballing by regularly asking the trial investigators involved whether they were aware of trials that were not yet included (final elicitation October 3, 2022).

Trials were eligible if they met the following criteria: 1) individual randomisation, 2) comparison of BCG to either placebo or no intervention, 3) population consists of adult healthcare workers, 4) COVID-19 and COVID-19 related hospitalisation are among the primary or secondary outcome measures.

### Trial inclusion criteria

Inclusion decisions were made on the study level. The decision to include a trial was based on (a) an external risk-of-bias assessment, (b) a specific type of expected homogeneity in effect sizes, and (c) an agreement on event definitions. These are described in detail below.

#### (a) Risk-of-bias assessment

Risk of bias was assessed at the study level using a modified version of the Cochrane risk-of-bias tool (Higgins, Altman, Gøtzsche, & al., 2011). The tool was modified to make it fit for assessing trial protocols, rather than publications of completed trials with results. Risk of bias was assessed for the following domains: random sequence generation, allocation concealment, blinding of participants, blinding of outcome assessment, method of outcome assessment and other bias. From the original risk of bias tool, we removed the domain selective reporting as there is no reporting of results in a protocol, and we changed the domain incomplete outcome data to method of outcome assessment. In this domain we assessed whether the method of collecting outcomes was appropriate and whether we expected that any outcomes could be missed by the researchers. Risk of bias was scored independently by two reviewers: one reviewer with a methodological background (AD) and one of two reviewers with a clinical background (KH and JW). All risk of bias domains (agreements and disagreements) were subsequently discussed within the review team.

During advisory board meetings, each trial’s risk-of-bias assessment was discussed, and advisory board members were invited to ask clarifying questions and to share their opinion. If there were concerns about possible biases and the data was already uploaded, the meta-trial statistician arranged a description of the data structure that was blinded to BCG vaccine allocation. The final decision for inclusion of a trial was made by the Steering committee.

#### (b) Expected homogeneity in effect sizes

The meta-analysis relies on a notion of qualitative trial homogeneity that is sufficient to make decisions during the pandemic. If evidence arises that BCG has a beneficial effect in the prevention of COVID-19, the analysis could serve as the leading source of information to start, stop or expand trials, and for countries to implement BCG vaccination in their population. Such evidence may also inform decisions in other countries and trial settings with slightly different population characteristics, historical BCG vaccination policies, different BCG strains, or different COVID-19 event definitions e.g. a trial based on PCR testing that informs decisions in trials based on antigen testing. As long as not enough data is available to do post-hoc subgroup analyses, a pandemic situation would require acting on what is available. In cases where it is possible to test differences between trials this would of course be recommended before acting on the evidence. However, the possibility of detailed subgroup analyses was not anticipated given the expected small effect sizes and large amounts of data needed to distinguish differences between small effects.

This rationale of decision making requires a specific argument of expected homogeneity in the effect sizes as a key criterium in the decision on trial inclusion, such that the meta-analysis could be the leading source of information. Trials were only included if we expected each of them to have an effect in the same direction (benefit, not harm) – if there would be benefit in one trial, we expect it in all trials. Moreover, trials were included if they could be expected to have an effect of a certain minimal size (if BCG has an effect at all). This connects to the use of fixed-effects (plural) meta-analysis discussed below. We follow Peto (1987) in this regard: “In performing overviews, we are not trying to provide exact quantitative estimates of percentage risk reductions in some precisely defined population of patients. We are simply trying to determine whether or not some type of treatment tested in a wide range of trials produces any effect […]”.

#### (c) Event definitions

##### Primary analysis

In the original statistical analysis plan that was shared with the participating trials, an event of COVID-19 infection was defined as *“Documented COVID-19 disease is defined as PCR-based detection of SARS-CoV-2 in a respiratory sample”*. During an advisory committee meeting on April 23^rd^, 2021 this was expanded to lung CTs and SARS-CoV-2 antigen positive, rapid point of care testing for current infection. The date of each event was set at the time when the test or scan were performed. For all trials having a positive test at randomisation (either PCR, serology, or otherwise) was an exclusion criterium for the trial itself (see detailed trial characteristics in section ***Detailed results***), or for data-extraction for the meta-analysis. Some trials (e.g. BR) also detected positive COVID-19 cases with serology during the course of the trial. These cases were not included in the COVID-19 event count in the primary analysis, but the participants that had a positive serology were also not deleted from the dataset, to not introduce bias. This means that they stayed in the risk set for COVID-19 infections detected by other means than serology, even though they were at different risk from participants without positive serology.

##### Secondary analysis

On December 1^st^, 2022 it was decided to perform a secondary analysis including trials that have the majority of events detected by SARS-CoV-2 serology defined as SARS-CoV-2 antibody positive/detected after infection, with the date of serology as the event time.

### Data extraction

Each trial had a designated data-uploader that also attended several Advisory board meetings and would become a meta-analysis co-author. IPD was extracted from each trial and uploaded to a secured cloud server in repeated uploads described in the ‘Working instruction for data-uploaders’, Statistical Analysis Plan and a webinar that explained the statistical methodology (ALL-IN-META-BCG-CORONA Replication Package). Data-uploaders were encouraged to check their data as it appeared in the dashboard (see Figure 2) based on a data processing tutorial (ALL-IN-META-BCG-CORONA Replication Package) and upload new data if available. The following variables were included: intervention randomised to (control or BCG), calendar date of randomisation, the stratification factor ‘hospital’ (e.g. “A”, “B”, “C”, etc) (see Appendix section ***Detailed results*** for the meaning of ‘hospital’ in the analysis of each trial), COVID-19 infection (yes/no) and calendar date the positive test or scan for COVID-19, COVID-19 related hospitalisation (yes/no) and calendar date of being hospitalised for COVID-19-related reasons, and calendar date of last follow-up. For patients still in follow-up at the time of data extraction, the last follow-up date was the calendar date of data extraction. The meta-analysis statisticians were in continuous contact with the data-uploaders to correct mistakes (dates before the COVID-19 pandemic, dates of randomisation later in time than dates of COVID-19 infection etc.).

### Participants at risk

Participants were considered at risk of COVID-19 infection and hospitalisation from the date of randomisation to the date of either a COVID-19 infection/hospitalisation, the end of follow-up, loss to follow-up or date of first COVID-19 specific vaccination. Therefore, follow-up time was censored at the date of first COVID-19 specific vaccination. Infections occurring after COVID-19 vaccination and reinfections were not included as events. Participants remained at risk of COVID-19 hospitalisation after a COVID-19 infection if this did not result in hospitalisation.

### Fixed-effects meta-analysis

The ALL-IN meta-analysis uses the *e*-values from the exact *e-*value logrank test (Ter Schure, Pérez-Ortiz, Ly, & Grünwald, 2022) and anytime-valid confidence intervals for the hazard ratio (HR) described in the ***Statistical Appendix*** that accompanies this paper.

The *e-*value analysis tests the *global null hypothesis* of no effect in all trials. This global null assumes a HR of 1 in all trials throughout their entire course. All events were analysed stratified by hospital within trial, by calculating hospital specific *e*-values and multiplying those into trial specific *e*-values, which are multiplied into the ALL-IN META *e*-values.

The approach to anytime-valid confidence intervals in this meta-analysis is the fixed-effects (plural) model following the logic of (Peto, 1987) describing a typical effect that is a weighted average of all the trials that contribute to the analysis (Rice, Higgins, & Lumley, 2018; Hedges & Vevea, 1998). The analysis assumes that the HR *can vary* from trial to trial, and is a two-stage analysis. A Cox proportional hazards model (maximum-likelihood) estimate was obtained for each trial, stratified by hospital (i.e. a single HR per trial that follows from evaluating all events with regard to the risk set in their hospital alone instead of the risk set in the full trial). These trial estimates were combined into a meta-analysis HR estimate using inverse-variance weighting. The event times are analysed in calendar time such that all participants within the same hospital at a given date are at the same risk, regardless of their own time since inclusion. More details about the exact scripts used are available in the Appendix section ***Detailed methods*** and supplementary material R code (ALL-IN-META-BCG-CORONA Replication Package).

The Kaplan-Meier curves that were promised in our Statistical Analysis Plan appeared difficult to interpret given the left-truncation of analysis in calendar time. The appearance of events is therefore presented as sequences of *e*-values and confidence intervals over time, with accompanying subplots indicating recruitment and follow-up period for each trial. The plot illustrating the sequence of confidence intervals is related to a single cumulative hazard plot for comparison.

### E-value analysis design: effect sizes of minimal clinical relevance and thresholds

An effect size of minimal clinical relevance was specified – arbitrarily – at HR = 0.8 for COVID-19 infections and at HR = 0.7 for COVID-19 hospitalisations. These corresponded well with the Food and Drug Administration recommendation to reject a null hypothesis Vaccine Efficacy of 0-30% (HR 0.7-1) in the COVID-19 pandemic, that was published soon after (FDA, Development and Licensure of Vaccines to Prevent COVID-19., 2020). In contrast to vaccines in development for COVID-19, the BCG vaccine was already widely available at a low price, and producing it at scale was considered possible. Hence the minimal relevant effect on reducing COVID-19 infections was kept at a smaller Vaccine Efficacy of 20% (HR = 0.8), while the effect on reducing hospitalisations was kept at 30% (HR = 0.7).

The main aim was to evaluate if the BCG vaccine was able to reduce severe disease and alleviate the burden on hospitals worldwide. A reduction in infections would rationally result in a reduction of hospitalisations as well, such that the former was considered as important, and that the power of that analysis would be larger with an expected higher event rate. Following this rationale, the two outcomes were set as co-primary outcomes and tested at the level α = 5%, with a Bonferroni correction spending 10% of 5% (0.5%) on infections and 90% of 5% (4.5%) on hospitalisations. This is an approach that loosely agrees with the FDA two-trial rule (two trials at α-level 5% (FDA, 1998, p. 3)) translating into an α = 0.25% meta-analysis (0.05*0.05 = 0.0025), two-sided. This was the α-level of the main analysis of interest (the analysis with the most power), that was a further restriction from two-sided, being the one-sided test for benefit on COVID-19 infections at level α = 0.25%.

Since statistical tests were performed for each side separately, a two-sided α of 0.5% for COVID-19 infections means 0.25% for each side and 4.5% for COVID-19 hospitalisations means 2.25% for each side. In terms of *e*-values this translates into our threshold for COVID-19 infections comparing the null of HR 1 to a smaller risk (left-sided test for benefit at HR of 0.8 or smaller) lies at 1/0.0025 = 400, and similarly for the *e-*value comparing the null to a larger risk (right-sided test for harm at HR of 1/0.8 or larger) at 400. For COVID-19 hospitalisations the threshold for the left-sided test (for benefit) and right-sided (for harm) comparing HR 1 to 0.7 and 1/0.7 lie at 1/0.0225 = 44.

### Design for early stopping for efficacy, not futility

The design decisions for the e-values – the effect sizes of minimal interest and α-levels stated above – were made by the Steering committee on May 29, 2020. This set a clear rule for what to watch out for in the dashboard, as illustrated by the dotted lines at 400 for COVID-19 infections and 44 for COVID-19 hospitalisations (shown in the demo dashboard in *Figure 2* and results in *Figure 4*).

No such threshold was decided for futility. A very early conclusion on futility was deemed unlikely, and a conclusion in the final follow-up phase of all trials would not prevent much wasted effort on further recruitment. As such we reported the *e-*values compared to the predetermined threshold for efficacy set at level *α* of 0.5% (infections) and 4.5% (hospitalisations), but we do not emphasise these same α-levels for the anytime-valid confidence intervals in a judgement of futility. Nevertheless, also without a threshold for futility, we can use confidence intervals to draw conclusions. If in all trials the HR is more extreme than a certain value, then it is very unlikely that such interesting extreme effect will fall outside of the interval. In infinite repeated use the chance that this happens for the anytime-valid 95%-confidence interval is at most 5%, no matter how long we keep updating (see Appendix ***Detailed methods***).

### Registration

The meta-analysis design was agreed on by the Steering Committee on May 29, 2020 and time*-*stamped in a logrank design object within R that is visible in the dashboard (Ter Schure J., ALL-IN-META-BCG-CORONA dashboard, 2020), see *Figure 2* on page *5*. Working Instructions for data-uploaders and the Statistical Analysis Plan (SAP) were made publicly available on the project website (Ter Schure, Ly, & Grünwald, 2020) on June 17, 2020 and were registered in the International prospective register of systematic reviews (PROSPERO: CRD42021213069) on February 11, 2021 (Van Werkhoven, et al., 2021). The SAP was updated into the version 2 available on the project website and in the Supplementary material (ALL-IN-META-BCG-CORONA Replication Package) accompanying this publication on September 19, 2022 and PROSPERO was updated on December 5, 2022.

## Results

### Trial inclusion

A total of 20 protocols were identified of trials of BCG for prevention of COVID-19 in healthcare workers. Of these, data from 6 trials were included in the current primary meta-analysis. A secondary analysis was defined that adds COVID-19 infections based on serology, and includes the AF trial as the 7nd trial. This process is described in Figure 3 and trial characteristics and summary data are described in Table 1 and Table 2. More trial details are provided in the Appendix section ***Detailed results***. Three protocols for risk-of-bias assessment were published, for the NL trial (Ten Doesschate, et al., 2020), the DK trial (Madsen, et al., 2020) and the BR trial (Junqueira-Kipnis, et al., 2020).

**Figure 3.**
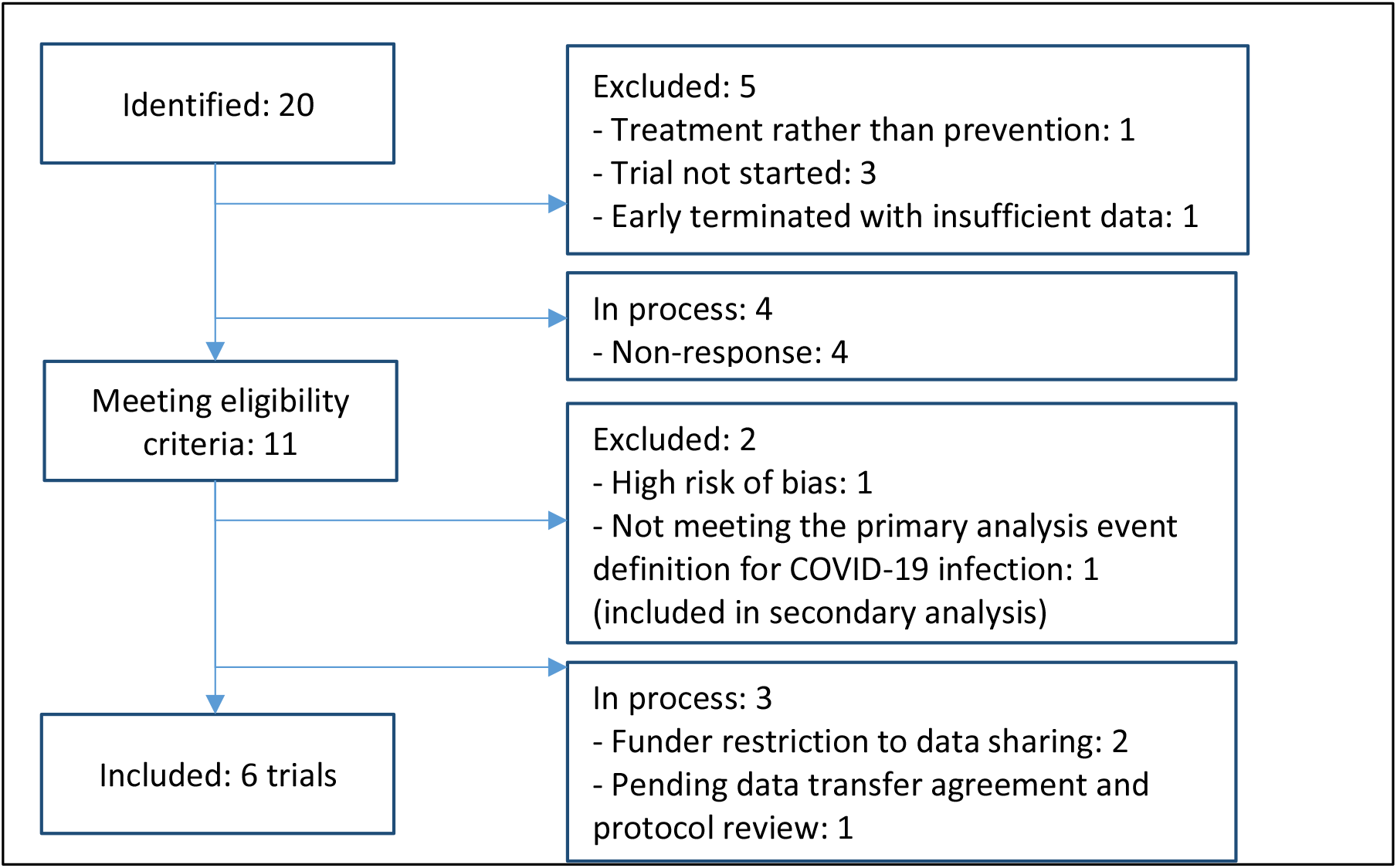
Flowchart of included trials

**Figure 4.**
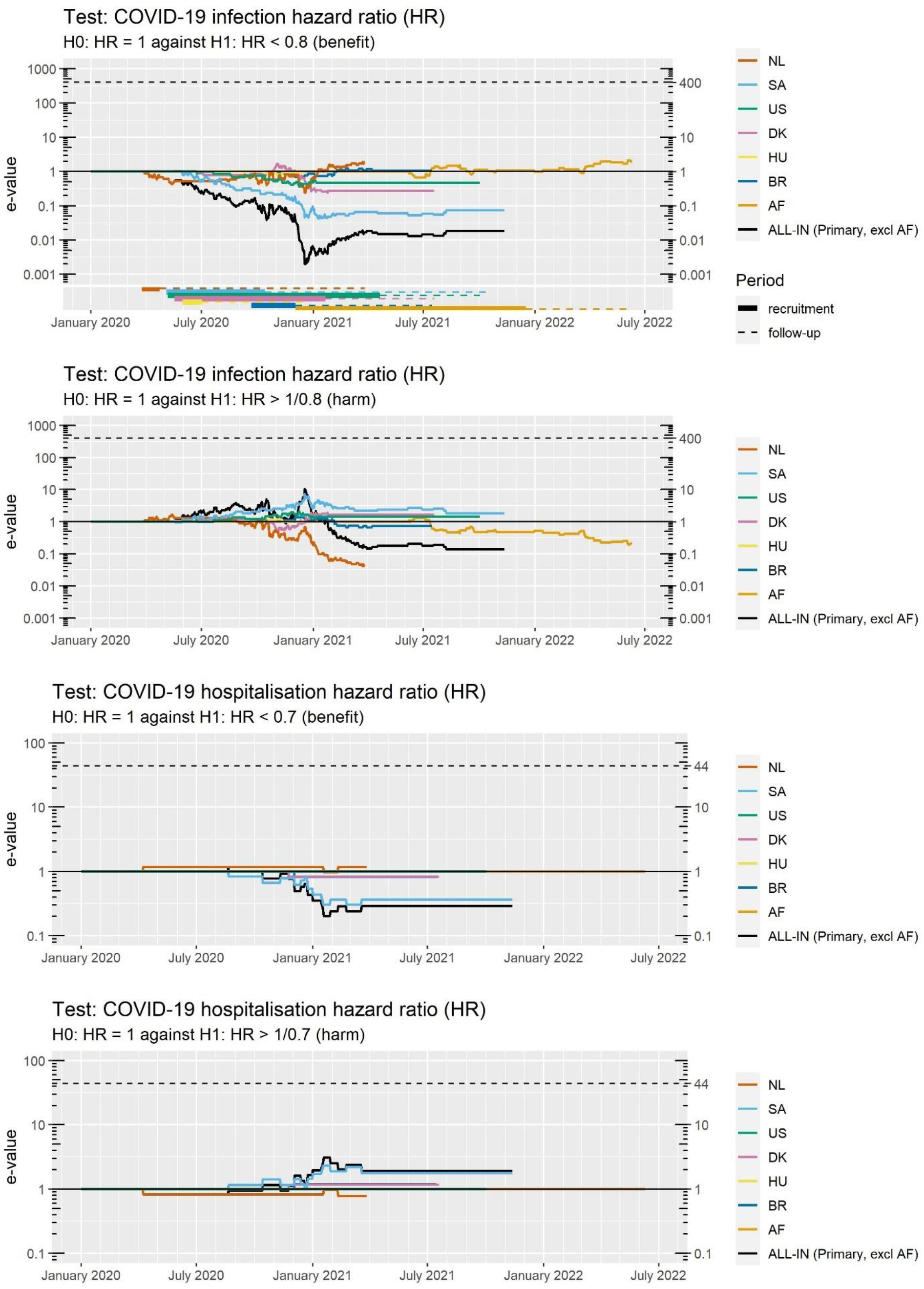
Exact logrank e-values for COVID-19 infections and hospitalisations with the Primary ALL-IN meta-analysis that excludes the AF trial. Note that the y-axis is logarithmic.

**Table 1.** Trial characteristics of included trials, with AF only included in the Secondary analysis

| **Trial** | **Definition event**<br>**COVID-19 infection** | **Intervention** | **Control** | **Prior BCG vaccination** |
| --- | --- | --- | --- | --- |
| **NL** | PCR and (a few) antigen | BCG Danish strain 1331 | Placebo | 17% self-reported |
| **SA** | PCR and antigen | BCG Danish strain 1331 | Placebo | BCG in National immunisation program; 50% had BCG scar |
| **US** | PCR and antigen | TICE® BCG | Placebo | 11% self-reported |
| **DK** | PCR and (very few) antigen | BCG Danish strain 1331 | Placebo | 53% self-reported; 49% had BCG scar |
| **HU** | PCR | BCG Brazilian Moreau substrain | Placebo | Compulsory BCG vaccination at birth since 1954 |
| **BR** | PCR | BCG Moscow 361-I | No vaccination | Restricted to individuals with a BCG scar |
| **AF\*** | (a few) PCR; serology at end of 6 month follow-up (96% of events, n=77) | BCG Danish strain 1331 | Placebo | Self-reporting unreliable due to many unknowns; 76% had BCG scar |

\* Guinea-Bissau/Mozambique

**Table 2.** Trial summary statistics

| **trial** | **Date first**<br>**randomised** | **Date last**<br>**follow up** | **Number**<br>**randomised** |  | **Person-weeks of**<br>**follow-up** |  | **COVID-19**<br>**infections** |  | **COVID-19**<br>**hospitalisations** |  |
| --- | --- | --- | --- | --- | --- | --- | --- | --- | --- | --- |
|  |  |  | BCG | control | BCG | control | BCG | control | BCG | control |
| **NL** | 25-Mar-2020 | 27-Mar-2021 | 747 | 749 | 33 523 | 33 006 | 96 | 110 | 1 | 2 |
| **SA** | 4-May-2020 | 18-Oct-2021 | 500 | 500 | 18 515 | 18 746 | 90 | 82 | 10 | 5 |
| **US** | 6-May-2020 | 2-Oct-2021 | 292 | 283 | 5 736 | 5 720 | 18 | 13 | 0 | 0 |
| **DK** | 18-May-2020 | 19-Jul-2021 | 610 | 611 | 11 466 | 11 386 | 36 | 27 | 1 | 0 |
| **HU** | 31-May-2020 | 18-Dec-2020 | 4 | 6 | 100 | 113 | 2 | 1 | 0 | 0 |
| **BR** | 21-Sep-2020 | 15-Jul-2021 | 64 | 67 | 1 807 | 1 949 | 9 | 11 | 1 | 0 |
| **AF\*** | 3-Dec-2020 | 10-Jun-2022 | 184 | 180 | 4 009 | 3 677 | 44 | 36 | 0 | 0 |
| **ALL-IN**<br>**Primary**<br>**(excl AF)** | 25-Mar-2020 | 18-Oct-2021 | 2 217 | 2 216 | 71 145 | 70 920 | 251 | 244 | 13 | 7 |
| **ALL-IN**<br>**Secondary**<br>**(incl AF)** | 25-Mar-2020 | 10-Jun-2022 | 2 401 | 2 396 | 75 155 | 74 597 | 295 | 280 | 13 | 7 |

\* Guinea-Bissau/Mozambique

**Table 3.** Exact logrank e-values for COVID-19 infections and hospitalisations

| trial | COVID-19 infections |  |  | COVID-19 hospitalisations |  |  |
| --- | --- | --- | --- | --- | --- | --- |
|  | Number of events | Exact e-value |  | Number of events | Exact e-value |  |
|  |  | Benefit | Harm |  | Benefit | Harm |
|  |  | H0: HR = 1<br>against<br>H1: HR < 0.8 | H0: HR = 1<br>against<br>H1: HR > 1/0.8 |  | H0: HR = 1<br>against<br>H1: HR < 0.7 | H0: HR = 1<br>against<br>H1: HR > 1/0.7 |
| NL | 206 | 1,884 | 0,041 | 3 | 1,167 | 0,780 |
| SA | 172 | 0,097 | 1,228 | 15 | 0,303 | 2,057 |
| US | 31 | 0,472 | 1,440 | 0 | 1 | 1 |
| DK | 63 | 0,274 | 1,673 | 1 | 0,828 | 1,170 |
| HU | 3 | 0,954 | 1,015 | 0 | 1 | 1 |
| BR | 20 | 1,053 | 0,741 | 1 | 0,821 | 1,180 |
| AF* | 80 | 1,982 | 0,212 | 0 | 1 | 1 |
| ALL-IN<br>Primary analysis<br>(excl AF) | 495 | 0,024 | 0,092 | 20 | 0,241 | 2,214 |
| ALL-IN<br>Secondary analysis<br>(incl AF) | 575 | 0,047 | 0,020 | 20 | 0,241 | 2,214 |

\* Guinea-Bissau/Mozambique

Following the rationale of a certain expected homogeneity in effect sizes (see Methods section ***Expected homogeneity in effect sizes***), it was decided prospectively to exclude trials in the elderly or more vulnerable population from the search for the same meta-analysis, because of the possibility that one of the two populations observed a small beneficial effect while the other observed a small effect of harm.

### Trials completed and ongoing

All trials presented here are completed, have locked their databases and shared their final data for the meta-analysis. Table 2 shows the date of last follow-up for each trial. The course of each trial is represented in *Figure 4* on page *14*, in a subplot indicating the period of recruitment and follow-up in each trial.

Figure 3 shows that three more trials are concluded and in process for their data to be added to this living systematic review. One has been in preparation for the data transfer agreement for 2,5 years, one cannot share IPD but is preparing aggregate data, and one is awaiting their own trial publication before joining the meta-analysis.

### Statistical results

The *e*-values show no evidence in favour of an effect of minimal clinically relevance (HR < 0.8) in comparison to the null (HR = 1) for COVID-19 infections and neither for COVID-19 hospitalisations (HR < 0.7 vs HR = 1). For the meta-analysis as a whole, we find an *e-*value of 0.023 for benefit for COVID-19 infections and an *e*-value of 0.241 for COVID-19 hospitalisations, indicating that the data is better supported by the null than the specific alternatives set for minimal clinical relevance. Overall, the results show that the *e-*values were never close to the thresholds at 400 and 44 for rejecting the null hypothesis, see *Figure 4*, and that the null hypothesis describes the data quite well, as indicated by the anytime-valid confidence intervals below. All hospital specific *e-*values are shown in the Appendix section ***Detailed results***.

#### COVID-19 infections

The primary meta-analysis estimate of the typical hazard ratio for COVID-19 infections was 1.02 with an anytime-valid 95%-confidence interval of (0.78-1.35). See the forest plot *Figure 5* and further details in *Table 4*. A secondary analysis including the AF trial with the majority of events confirmed by serology is shown in the forest plot in Figure 6. The primary analysis as well as the secondary analysis strongly suggest that the planned addition of events from trials not yet included in the meta-analysis – which is allowed in our anytime-valid approach – is highly likely to exclude the hypothesis that there is a minimal effect size of 0.8. Since this was our pre*-*specified minimum clinically relevant effect size, it would complete the meta-analysis with a futility conclusion, at the *α* = 5% level.

**Figure 5.**
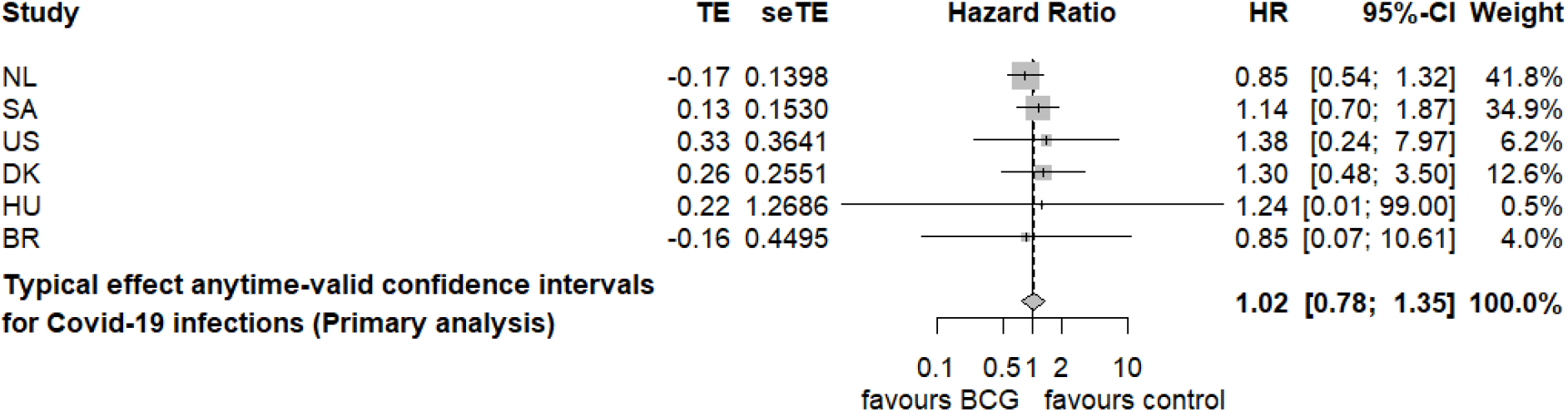
Primary analysis (excl AF) forest plot for the fixed-effects (plural) meta-analysis model

**Table 4.** Estimates for COVID-19 infections

| COVID-19 infections |  |  |  |  |  |  |
| --- | --- | --- | --- | --- | --- | --- |
| Trial | Number of events | Hazard Ratio (HR) |  |  |  |  |
|  |  | Maximum likelihood estimator Cox model stratified by hospital<br>Typical effect size | Anytime-valid 95% CI |  | Anytime-valid 99.5% CI |  |
|  |  |  | lower | upper | lower | upper |
| NL | 206 | 0,85 | 0,54 | 1,32 | 0,48 | 1,49 |
| SA | 172 | 1,14 | 0,70 | 1,87 | 0,60 | 2,16 |
| US | 31 | 1,38 | 0,24 | 7,97 | 0,14 | 13,84 |
| DK | 63 | 1,30 | 0,48 | 3,50 | 0,36 | 4,73 |
| HU | 3 | 1,24 | <0,01 | >99 | <0,01 | >99 |
| BR | 20 | 0,85 | 0,07 | 10,61 | 0,03 | 23,75 |
| AF* | 80 | 0,76 | 0,32 | 1,81 | 0,25 | 2,35 |
| ALL-IN Primary (excl AF) | 495 | 1,02 | 0,78 | 1,35 | 0,73 | 1,45 |
| Secondary (incl AF) | 575 | 0.98 | 0,76 | 1,27 | 0,71 | 1,36 |

\* AF: Guinea-Bissau/Mozambique

**Table 5.** Detailed characteristics NL trial

| Characteristic | Description |
| --- | --- |
| Trial abbreviation | NL |
| Country | The Netherlands |
| Authors representing trial | C.H. (Henri) van Werkhoven (data-uploader), Marc M.J. Bonten (PI) |
| Trial Registry | ClinicalTrials.gov: [NCT04328441](https://clinicaltrials.gov/ct2/show/study/NCT04328441) |
| Protocol publication | Ten Doesschate et al. *Trials*. 2020. DOI: [10.1186/s13063-020-04389-w](https://doi.org/10.1186/s13063-020-04389-w) |
| Results publication | Ten Doesschate et al. *Clin Microbiol Infect*. 2022. DOI: [10.1016/j.cmi.2022.04.009](https://doi.org/10.1016/j.cmi.2022.04.009) |
| Inclusion criteria | Adult ($\geq 18$ years)<br>Hospital personnel (expected to) taking care for patients with SARS-CoV-2 infection |
| Exclusion criteria | Known allergy to (components of) the BCG vaccine or serious adverse events to prior BCG administration<br>Known active or latent *Mycobacterium tuberculosis* or with another mycobacterial species. A history with- or a suspicion of *M. tuberculosis* infection.<br>Fever ($>38$ C) within the past 24 hours<br>Pregnancy<br>Suspicion of active viral or bacterial infection<br>Vaccination in the past 4 weeks or expected vaccination during the study period, independent of the type of vaccination.<br>Severely immunocompromised subjects. This exclusion category comprises: a) subjects with known infection by the human immunodeficiency virus (HIV-1); b) neutropenic subjects with less than 500 neutrophils/mm<sup>3</sup>; c) subjects with solid organ transplantation; d) subjects with bone marrow transplantation; e) subjects under chemotherapy; f) subjects with primary immunodeficiency; g) severe lymphopenia with less than 400 lymphocytes/mm<sup>3</sup>; h) treatment with any anti- cytokine therapies. i) treatment with oral or intravenous steroids defined as daily doses of 10mg prednisone or equivalent for longer than 3 months, or probable use of oral or intravenous steroids in the following four weeks<br>Active solid or non-solid malignancy or lymphoma within the prior two years<br>Direct involvement in the design or the execution of the BCG-CORONA study<br>Expected absence from work of $\geq 4$ of the following 12 weeks due to any reason (holidays, maternity leave, retirement, planned surgery etc)<br>Employed by the hospital $< 22$ hours per week<br>Not in possession of a smartphone |
| History of BCG vaccination at enrolment | Not restricted; 17% reported prior BCG vaccination in both groups |
| Description of Intervention | BCG Vaccine SSI [Statens Serum Vaccin Institut] Danish strain 1331, intradermal injection |
| Control group intervention | Placebo: intradermal injection of 0.1ml 0.9% NaCl, which is the same amount, and has the same color as the resuspended BCG vaccine |
| Outcome definition COVID-19 infection | Positive SARS-CoV-2 PCR or rapid antigen test |
| Type of randomisation | 1:1 by computer-generated randomisation in random blocks of 2, 4, or 6 sequences stratified by hospital |
| Definition of hospital as used in the analysis | Hospital of occupation, also used as stratum for randomisation |

**Table 6.** Detailed characteristics SA trial

| Characteristic | Description |
| --- | --- |
| Trial abbreviation | SA |
| Authors representing trial | Gerben van den Hoogen (data-uploader), Caryn M. Upton (PI) |
| Country | South-Africa |
| Trial Registry | ClinicalTrials.gov: [NCT04379336](https://clinicaltrials.gov/ct2/show/study/NCT04379336) |
| Protocol publication |  |
| Results publication | Upton et al. *eClinicalMedicine* 2022. DOI: [10.1016/j.eclinm.2022.101414](https://doi.org/10.1016/j.eclinm.2022.101414) |
| Inclusion criteria | • Adult $\geq 18$ years<br>• HCW and other frontline staff deemed at risk of exposure to SARS-CoV-2 as the COVID-19 epidemic emerges.<br>• NOTE: Frontline workers, as a general guide, are professionals or volunteers that in the course of the epidemic are deemed at increased risk of exposure.<br>• Ability and willingness to provide informed consent.<br>• Can be reached by mobile phone for follow-up |
| Exclusion criteria | • Known allergy to (components of) the BCG vaccine or serious reaction to prior BCG administration.<br>• Known active tuberculosis or any other active or uncontrolled condition that, in the opinion of the investigator or designee, makes participation unsafe or makes it difficult to collect follow-up data over the study period.<br>• HIV-1 infection<br>• NOTE: If evidence of recent HIV negative test (within the last year) is not available, rapid point-of-care testing will be undertaken as part of screening with a separate informed consent process.<br>• Symptoms of respiratory tract infection which, in the opinion of the investigator or designee, is likely to interfere with the objectives of the study.<br>• Known current or previous infection with SARS-CoV-2.<br>• Known medical history of any of the following immunocompromised states:<br>○ Neutropenia (less than 500 neutrophils/mm<sup>3</sup>)<br>○ Lymphopenia (less than 400 lymphocytes/mm<sup>3</sup>)<br>○ Solid organ or bone marrow transplantation<br>○ Primary immunodeficiency<br>○ Active solid or non-solid malignancy or lymphoma within the prior two years<br>○ Pregnancy and breastfeeding<br>• Current treatment with the following medications:<br>○ Chemotherapy<br>○ Anti-cytokine therapies<br>○ Current treatment with oral or intravenous steroids defined as daily doses of 10mg prednisone or equivalent for longer than 3 months<br>○ Any experimental, unproven treatment against SARS-CoV-2 infection or COVID-19 including but not limited to chloroquine, hydroxychloroquine, remdesivir, lopinavir/ritonavir and interferon beta- 1a. |
| History of BCG vaccination at enrolment | Not restricted: BCG vaccination at age of 6 weeks is part of national vaccination program. BCG scar present: 259 (51.8%) in the placebo arm, 237 (47.4%) in the BCG arm |
| Description of Intervention | BCG Vaccine SSI [Statens Serum Vaccin Institut] Danish strain 1331, intradermal injection |
| Control group intervention | Placebo: 0.1ml 0.9% NaCl, which is the same volume and has the same colour as the suspended BCG vaccine |
| Outcome definition COVID-19 infection | Positive SARS-CoV-2 PCR or rapid antigen test |
| Type of randomisation | No stratification |
| Definition of hospital as used in the analysis | Geographical stratum code |

**Table 7.** Detailed characteristics US trial

| Characteristic | Description |
| --- | --- |
| Trial abbreviation | US |
| Authors representing trial | Jose Euberto Mendez-Reyes (data-uploader), Jeffrey D. Cirillo (PI) |
| Country | The United States |
| Trial Registry | ClinicalTrials.gov: [NCT04348370](https://clinicaltrials.gov/ct2/show/study/NCT04348370) |
| Protocol publication |  |
| Results publication |  |
| Inclusion criteria | • Adult ($\geq 18$ years)<br>• High risk individual including:<br>• Health Care Workers (HCW) Personnel working in a healthcare setting, at a hospital, medical centre or clinic (veterinary, dental, ophthalmology), or first responders (paramedics, firefighters, or law enforcement).<br>○ High risk for severe disease including elderly and those with comorbidities including obesity (BMI > 25), elderly (age > 65 years), hypertension, diabetes, reactive airway disease, smokers<br>○ Individuals at increased risk of infection because of decreased ability to limit exposure including racial and ethnic minorities, teachers, police, restaurant wait-staff, delivery personnel, grocery store and retail workers |
| Exclusion criteria | • Known allergy to (components of) the BCG vaccine or serious adverse events to prior BCG administration<br>• Known active or latent Mycobacterium tuberculosis or with another mycobacterial species. A history with- or a suspicion of M. tuberculosis infection.<br>• Fever (>38 C) within the past 24 hours<br>• Pregnancy or planning pregnancy within 30 days of study enrolment<br>• Breastfeeding<br>• Suspicion of active viral or bacterial infection<br>• Any Immunocompromised subjects. This exclusion category comprises: a) subjects with known infection by the human immunodeficiency virus (HIV-1); b) subjects with known neutropenic with less than 1500 neutrophils/mm<sup>3</sup>; c) subjects with solid organ transplantation; d) subjects with bone marrow transplantation; e) subjects under chemotherapy; f) subjects with primary immunodeficiency; g) known severe lymphopenia with less than 400 lymphocytes/mm<sup>3</sup>; h) treatment with any anti- cytokine therapies; i) treatment with oral or intravenous steroids defined as daily doses of 10mg prednisone or equivalent for longer than 3 months; j) taking immunosuppressants<br>• Living with someone who is immunosuppressed or taking immunosuppressive drugs<br>• Previous documented infection with COVID19<br>• Active solid or non-solid malignancy or lymphoma within the prior two years<br>• Direct involvement in the design or the execution of the study<br>• Not in possession and/or access to use of a smartphone, tablet or computer<br>• Inability to keep the vaccine site covered in the case of a draining pustule. |
| History of BCG vaccination at enrolment | Not excluded; 11% reported prior BCG vaccination in both groups |
| Description of Intervention | TICE® BCG (for intravesical use) BCG LIVE strain of the BCG (Merck), intradermal injection |
| Control group intervention | Placebo: Intradermal injection of 0.1 mL 0.9% NaCl, which is the same amount and color as the intervention |
| Outcome definition COVID-19 infection | Positive SARS-CoV-2 PCR or rapid antigen test |
| Type of randomisation | Stratified by study site |
| Definition of hospital as used in the analysis | Hospital of occupation with hospitals merged into one for each site that was also used as stratum for randomisation |

**Table 8.**
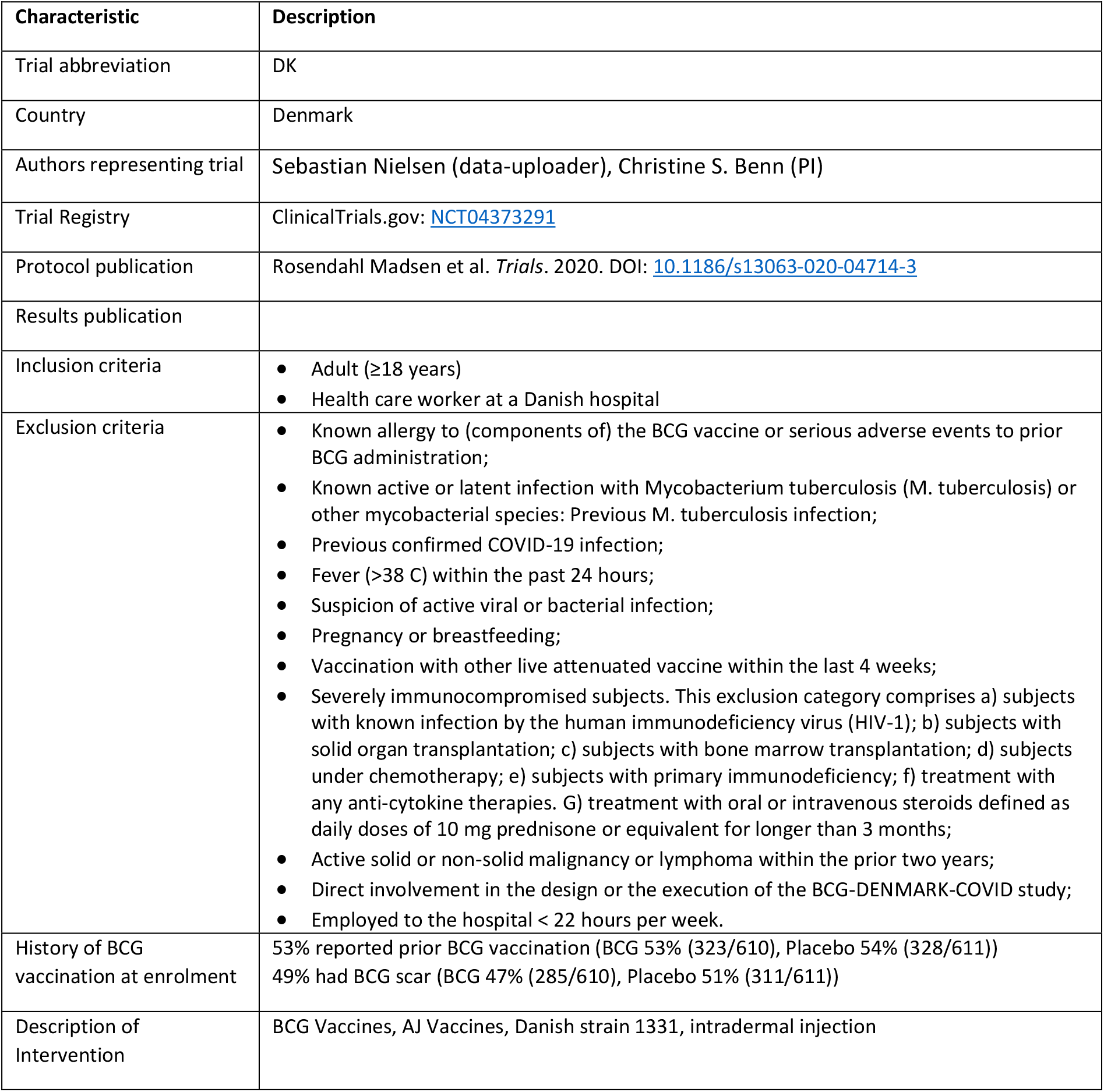
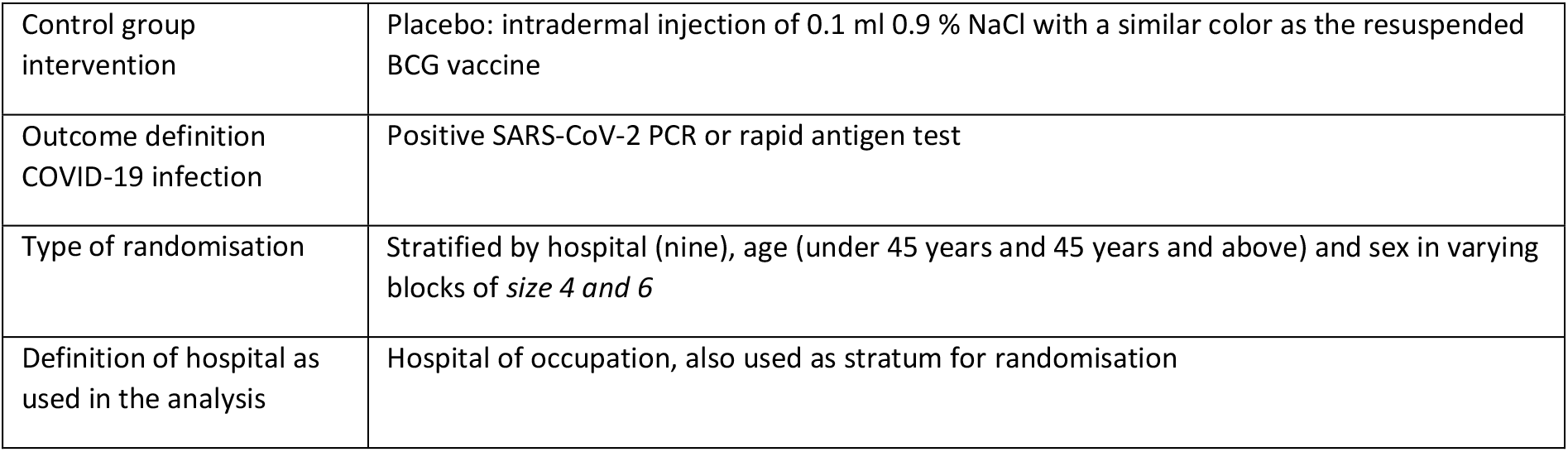
Detailed characteristics DK trial

**Table 9.** Detailed characteristics HU trial

| Characteristic | Description |
| --- | --- |
| Trial abbreviation | HU |
| Authors representing trial | Judit Moldvay (PI) |
| Country | Hungary |
| Trial Registry | EU Clinical Trials Register: [2020-001783-28](https://clinicaltrials.gov/ct2/show/study?term=2020-001783-28) |
| Protocol publication |  |
| Results publication |  |
| Inclusion criteria | • Adult (≥18 years)<br>• HCW taking care for patients with SARS-CoV-2 infection<br>• Signature of information consent form |
| Exclusion criteria | • Known allergy to (components of) the BCG vaccine or serious adverse events to prior BCG administration<br>• Quantiferon positivity. Known active or latent Mycobacterium tuberculosis or with another mycobacterial species. A history with- or a suspicion of M. tuberculosis infection.<br>• Positive COVID-19 test<br>• Fever (>38 C) within the past 72 hours<br>• Suspicion of active viral or bacterial infection<br>• CRP: >20 mg/l<br>• Not controlled diabetes melitus<br>• Inflammatory immune disease (eg SLE [systemic lupus erythematosus], RA [rheumatoid arthritis], polymyositis, systemic sclerosis, Sjögren's syndrome, systemic vasculitis, IBD [inflammatory bowel disease], SM [multiple sclerosis])<br>• Vaccination in the past 4 weeks or expected vaccination during the study period, independent of the type of vaccination.<br>• Severely immunocompromised subjects. This exclusion category comprises: a) subjects with known infection by the human immunodeficiency virus (HIV-1); b) neutropenic subjects with less than 500 neutrophils/mm<sup>3</sup>; c) severe lymphopenia with less than 400 lymphocytes/mm<sup>3</sup>; d) subjects with solid organ transplantation; e) subjects with bone marrow transplantation; f) subjects under chemotherapy; g) subjects with primary immunodeficiency; h) those with IgG levels below the lower limit of normal; i) treatment of synthetic (eg methotrexate, cyclophosphamide) or targeted (biological therapies eg TNF-blockers, IL-6 blockers, targeted synthetic disease modifiers (eg tofacitinib, bari-citinib) within one year; j) treatment with oral or intravenous steroids defined as daily doses of 5mg prednisone or equivalent for longer than 3 months, or probable use of oral or intravenous steroids in the following 12 weeks<br>• Active solid or non-solid malignancy or lymphoma within the prior 5 years<br>• Pregnancy<br>• Direct involvement in the design or the execution of the BACH study |
|  | Expected absence from work of $\geq 10$ days of the following 12 weeks due to any reason (holidays, maternity leave, retirement etc)<br>Employed by the hospital < 10 hours per week |
| History of BCG vaccination at enrolment | BCG vaccination at birth is compulsory since 1954 |
| Description of Intervention | Biomed Lublin, Brazilian Moreau substrain, intradermal injection |
| Control group intervention | Placebo: intradermal injection of 0.1ml NATRIUM CHLORATUM TEVA 0.9% suspension injection (OGYI-T-9776/03) which has the same color and same amount as the resuspended BCG vaccine |
| Outcome definition COVID-19 infection | Positive SARS-CoV-2 PCR or rapid antigen test |
| Type of randomisation | Randomisation was central, computer-generated, with stratification of patients by hospital into groups of 2, 4, and 6. |
| Definition of hospital as used in the analysis | Single hospital (single-centre trial) |

**Table 10.**
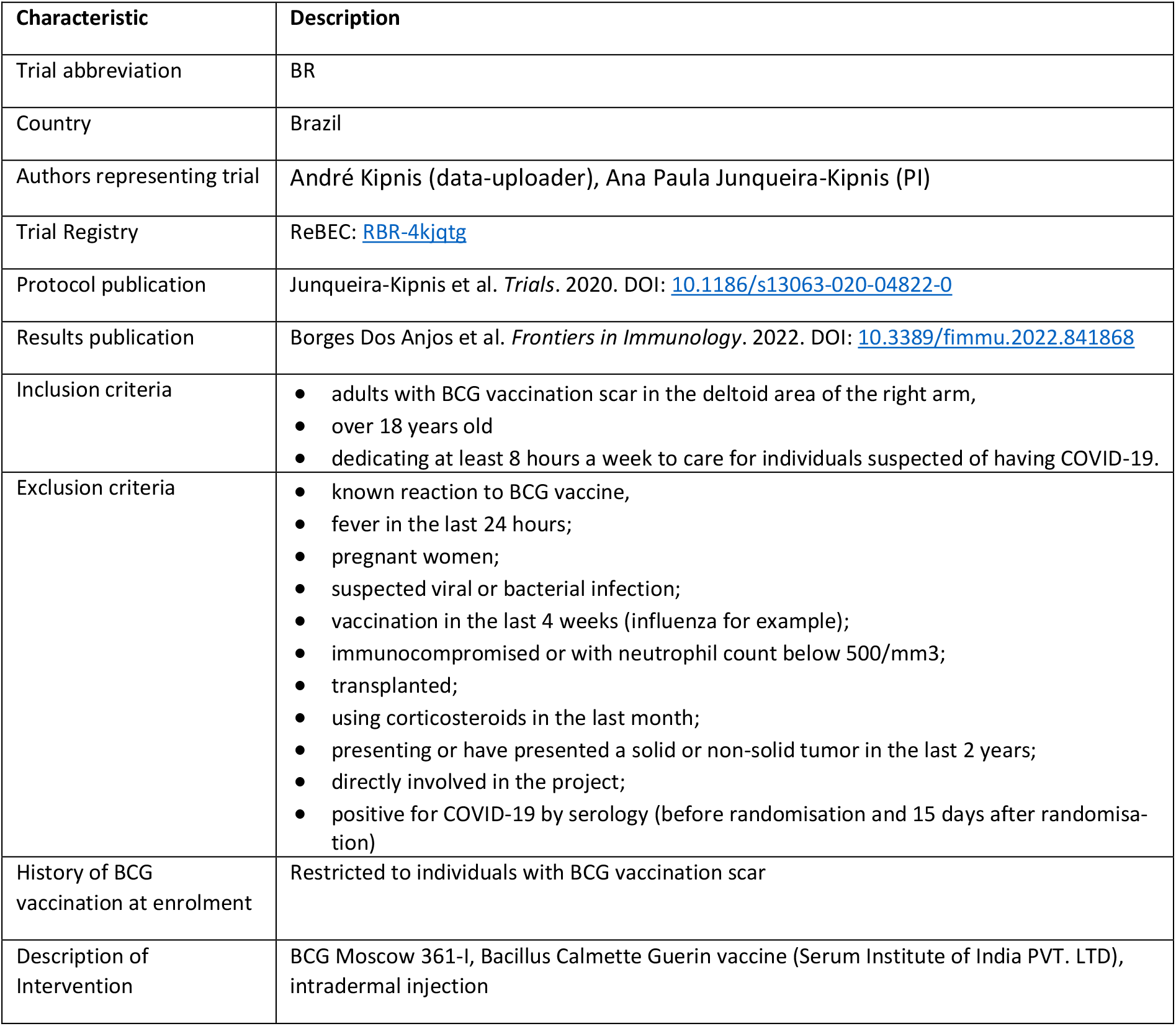
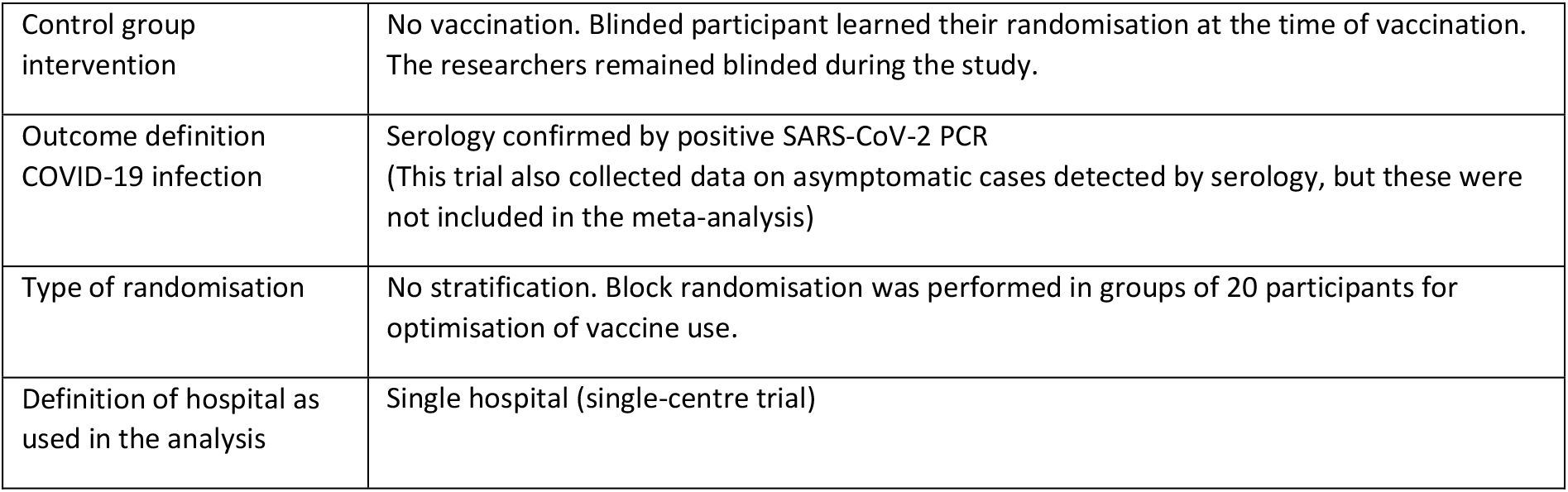
Detailed characteristics BR trial

**Table 11.** Detailed characteristics AF trial

| Characteristic | Description |
| --- | --- |
| Trial abbreviation | AF |
| Country | Guinea-Bissau and Mozambique |
| Authors representing trial | Sebastian Nielsen (data-uploader), Inês Fronteira (PI) |
| Trial Registry | ClinicalTrials.gov: [NCT04641858](https://clinicaltrials.gov/ct2/show/study/NCT04641858) |
| Protocol publication |  |
| Results publication |  |
| Inclusion criteria | Health care worker;<br>age $\geq 18$ years. |
| Exclusion criteria | known allergy to (components of) BCG or serious adverse events to prior BCG administration;<br>known previous, active or latent infection with Mycobacterium tuberculosis or other mycobacterial species<br>No testing for tuberculosis infections was performed, but a score was calculated using the TB score and participants scoring higher than 2 were excluded from the trial.<br>fever ($>38$ C) within past 24 hours;<br>previous confirmed COVID-19 (positive test - PCR or antibody);<br>negative serology test of COVID-19 antibodies at time of inclusion before the introduction of COVID-19 vaccines. (After introduction of COVID-19 vaccines participants were tested but not excluded in the main trial based on the results of the serology test.<br>after the introduction of COVID -19 vaccines, participants were not included between doses of COVID-19 vaccines and only 2 weeks after having received the second dose or first dose of Johnson&Johnson. Or if they anticipated receiving a COVID-19 vaccine within the following 2 weeks.<br>suspicion of active viral or bacterial infection.)<br>severely immunocompromised subjects<br>self-reported HIV infection (in Guinea-Bissau all participants were also HIV-tested (quick test))<br>self-reported pregnancy;<br>active solid or non-solid malignancy or lymphoma within the prior two years;<br>contraindications for live attenuated vaccine administration.<br>not having a mobile phone.<br>vaccination with other live attenuated vaccine within the last 4 weeks |
| History of BCG vaccination at enrolment | Reported BCG vaccination unreliable since many reported unknown<br>76% had BCG scar (BCG 78% (143/184), Placebo 74% (134/180)) |
| Description of Intervention | BCG Vaccines, AJ Vaccines, Danish strain 1331, intradermal injection |
| Control group intervention | Placebo: Intradermal injection of standard 0.1 ml saline solution (NaCl 0.9%) |
| Outcome definition COVID-19 infection | (a few) PCR; serology at end of 6 month follow-up (96% of events, n=77) |
| Type of randomisation | Stratified by country, profession (doctor, nurse, other) and sex in varying blocks of size 4 and 6 |
| Definition of hospital as used in the analysis | Country, also used as stratum for randomisation |

**Table 12.** Full summary statistics per hospital within trial

| trial | hospital | Date first randomised | Date last follow up | Number randomised to BCG | Number randomised to control | Person-weeks of follow up BCG | Person-weeks of follow up control | Number of COVID-19 infections BCG | Number of COVID-19 infections control | Number of COVID-19 hospitalisations BCG | Number of COVID-19 hospitalisations control |
| --- | --- | --- | --- | --- | --- | --- | --- | --- | --- | --- | --- |
| NL | A | 1-Apr-2020 | 27-Mar-2021 | 38 | 37 | 1 559 | 1 492 | 8 | 6 | 0 | 0 |
|  | B | 31-Mar-2020 | 27-Mar-2021 | 21 | 21 | 927 | 927 | 4 | 3 | 0 | 1 |
|  | C | 26-Mar-2020 | 27-Mar-2021 | 48 | 49 | 2 073 | 2 009 | 3 | 11 | 0 | 0 |
|  | D | 27-Mar-2020 | 27-Mar-2021 | 25 | 25 | 1 129 | 1 104 | 5 | 3 | 0 | 0 |
|  | E | 30-Mar-2020 | 27-Mar-2021 | 28 | 28 | 1 283 | 1 226 | 1 | 5 | 0 | 0 |
|  | F | 26-Mar-2020 | 27-Mar-2021 | 158 | 158 | 7 060 | 6 888 | 24 | 30 | 1 | 0 |
|  | G | 25-Mar-2020 | 27-Mar-2021 | 208 | 208 | 9 501 | 9 244 | 18 | 21 | 0 | 0 |
|  | H | 30-Mar-2020 | 27-Mar-2021 | 29 | 34 | 1 318 | 1 587 | 2 | 8 | 0 | 0 |
|  | I | 25-Mar-2020 | 27-Mar-2021 | 192 | 189 | 8 673 | 8 529 | 31 | 23 | 0 | 1 |
|  | total | 25-Mar-2020 | 27-Mar-2021 | 747 | 749 | 33 523 | 33 006 | 96 | 110 | 1 | 2 |
| SA | A | 4-May-20 | 15-Oct-21 | 463 | 449 | 17 127 | 16 757 | 81 | 73 | 8 | 4 |
|  | B | 4-May-20 | 18-Oct-21 | 37 | 51 | 1 388 | 1 989 | 9 | 9 | 2 | 1 |
|  | total | 4-May-20 | 18-Oct-21 | 500 | 500 | 18 515 | 18 746 | 90 | 82 | 10 | 5 |
| US | A | 2-Jun-2020 | 10-May-2021 | 81 | 85 | 1 543 | 1 627 | 3 | 4 | 0 | 0 |
|  | B | 11-Jun-2020 | 10-May-2021 | 45 | 48 | 923 | 993 | 1 | 3 | 0 | 0 |
|  | C | 6-May-2020 | 2-Oct-2021 | 166 | 150 | 3 270 | 3 100 | 14 | 6 | 0 | 0 |
|  | total | 6-May-2020 | 2-Oct-2021 | 292 | 283 | 5 736 | 5 720 | 18 | 13 | 0 | 0 |
| DK | A | 18-May-2020 | 7-Dec-2020 | 46 | 45 | 1 158 | 1 155 | 2 | 1 | 0 | 0 |
|  | B | 2-Jun-2020 | 10-May-2021 | 76 | 77 | 1 413 | 1 419 | 2 | 3 | 0 | 0 |
|  | C | 20-May-2020 | 7-Jun-2021 | 157 | 154 | 3 265 | 3 142 | 14 | 10 | 0 | 0 |
|  | D | 11-Jun-2020 | 1-Mar-2021 | 43 | 44 | 1 036 | 1 003 | 1 | 3 | 0 | 0 |
|  | E | 9-Jun-2020 | 14-Dec-2020 | 24 | 24 | 614 | 604 | 0 | 1 | 0 | 0 |
|  | F | 22-Jun-2020 | 22-Feb-2021 | 51 | 50 | 1 231 | 1 219 | 6 | 2 | 0 | 0 |
|  | G | 8-Sep-2020 | 17-May-2021 | 66 | 66 | 945 | 978 | 3 | 0 | 0 | 0 |
|  | H | 17-Aug-2020 | 19-Jul-2021 | 54 | 56 | 744 | 802 | 2 | 2 | 1 | 0 |
|  | I | 14-Sep-2020 | 21-Jun-2021 | 93 | 95 | 1 060 | 1 066 | 6 | 5 | 0 | 0 |
|  | total | 18-May-2020 | 19-Jul-2021 | 610 | 611 | 11 466 | 11 386 | 36 | 27 | 1 | 0 |
| HU | total | 31-May-2020 | 18-Dec-2020 | 4 | 6 | 100 | 113 | 2 | 1 | 0 | 0 |
| BR | total | 21-Sep-2020 | 15-Jul-2021 | 64 | 67 | 1 807 | 1 949 | 9 | 11 | 1 | 0 |
| AF | A | 3-Dec-2020 | 10-Jun-2022 | 167 | 156 | 3 679 | 3 413 | 38 | 34 | 0 | 0 |
|  | B | 29-Apr-2021 | 10-Jun-2022 | 17 | 24 | 330 | 263 | 6 | 2 | 0 | 0 |
|  | total | 3-Dec-2020 | 10-Jun-2022 | 184 | 180 | 4 009 | 3 677 | 44 | 36 | 0 | 0 |
| ALL-IN Primary |  | 25-Mar-20 | 18-Oct-21 | 2 217 | 2 216 | 71 145 | 70 920 | 251 | 244 | 13 | 7 |
| ALL-IN Secondary |  | 25-Mar-20 | 10-Jun-22 | 2 401 | 2 396 | 75 155 | 74 597 | 295 | 280 | 13 | 7 |

**Figure 6.**
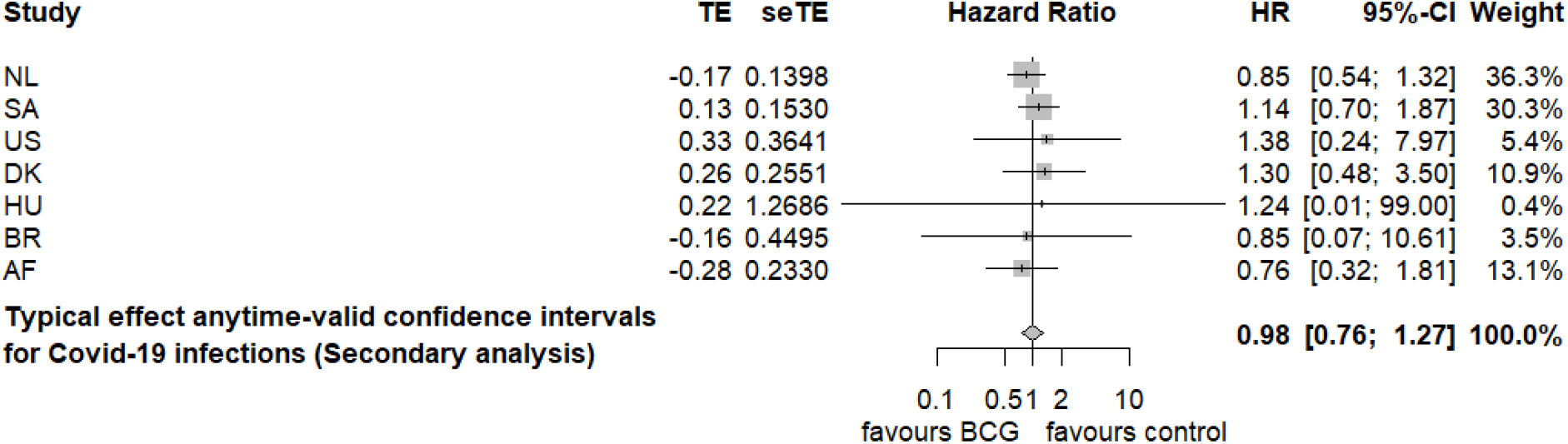
Secondary analysis (incl AF) forest plot for the fixed-effects (plural) meta-analysis model

#### COVID-19 hospitalisations

For events of COVID-19 hospitalisations only the NL, DK, BR and SA trials contribute events. However, for NL, DK and BR it is not possible to obtain a useful trial-specific confidence interval due to limited data; the estimation procedure did not converge (so we have no maximum likelihood estimate) and the intervals for the HR range from 0 to infinity. For SA, the maximum likelihood estimator in the Cox model stratified by hospital was 2.11 with (0.17-26.73) as its anytime-valid 95%-confidence interval. Because we have no estimate per trial, we cannot inverse-variance weigh these estimates to produce a meta-analysis estimate, as described in the ***Methods*** section and performed for COVID-19 infections. We can however, opt for a one-stage non-stratified approach and combine all data together in one dataset and analyse it stratified by trial (but not stratified by hospital). This achieves a maximum likelihood estimator of the Cox model that is still quite uninformative of 1.88 with (0.26, 13.40) as its anytime-valid 95%-confidence interval. Note that in contrast to the meta-analysis on COVID-19 infections, this analysis does assume that all strata (trials in this case) share a single HR, and does not stratify the baseline risk by hospital.

## Discussion

In this prospective and living IPD ALL-IN meta-analysis of completed and ongoing trials no effectiveness of BCG in reducing COVID-19 infection was observed. The precision of effect estimates is high and ‘almost’ excludes the minimal effect that was pre*-*specified to be of interest: the anytime-valid confidence interval lower end is 0.78 and the predetermined minimal relevant effect size was set at 0.8. For COVID-19 related hospitalisation, the limited number of events precluded a firm conclusion. This endpoint is rare in healthcare workers, especially with the less pathogenic SARS-CoV-2 Omicron variants circulating today and the availability of SARS-CoV-2 targeted vaccines.

*In vitro* and experimental studies demonstrated that BCG vaccination induces non-specific changes in the innate immune system that last for months (Netea, Domínguez-Andrés, Barreiro, & others, 2020). A decreased incidence of respiratory infections in adults after receiving BCG has been demonstrated by several small trials conducted before the SARS-CoV-2 pandemic (Datau, Sultana, Mandang, & others, 2011; Nemes, Geldenhuys, Rozot, & others, 2018; Giamarellos-Bourboulis, Tsilika, S, & al., 2020). The present results indicate that no such clear effect exists for COVID-19. Most of the previously published trials of BCG against COVID-19 in healthcare workers are included in the present analysis, with a trial from France in preparation. A global trial coordinated from Australia and another trial from Poland are not yet included due to restrictions by the funder. Results published from the Polish trial are in line with the meta-analysis (Czajka, et al., 2022). We did not include in the current meta-analysis trials performed in older adults, one small trial from Greece shows a reduction of the COVID-19 incidence in BCG-vaccinated individuals, while two larger trials from the Netherlands do not demonstrate an effect (Tsilika, et al., 2022; Ten Doesschate, et al., 2022; Koekenbier, 2021).. Also, a trial in type 1 diabetics found a strong protective effect of having received multiple doses of BCG within the last years (Faustman, et al., 2022). Why BCG vaccination would be protective against other respiratory tract infections but not SARS-CoV-2 remains a topic for further research. A recent experimental study demonstrating strong protection induced by BCG against influenza, but not COVID-19, suggested that important immunological and pathophysiological differences between the two infections may explain this observation (Kaufmann, et al., 2022). Noteworthy, a recent meta-analysis of all the published BCG-COVID-19 trials that reported deaths within the trials showed that BCG was associated with 39% (1-62%) reduction in all-cause mortality (Aaby, Netea, & Benn, 2022).

Our study was prospectively planned and analyses were designed before any trial data was available. This, together with the use of *e-*values and anytime-valid confidence intervals, is an important strength of our study, as it controls the type*-*I error rate even if new data is added in future updates of this analysis. In fact, the use of *e-*values was employed to allow for the use of interim meta-analysis results at any time for putative strategic decisions such as the initiation of a new trial, early termination, or extension of follow-up. Unfortunately, due to the delayed availability of data for the meta-analysis, the interim meta-analysis results were never used for such strategic decisions. In hindsight, the evidence against the hypothesis of superiority against COVID-19 infection hardly changed after 2020 (see Figure 7). Investigators might have decided to stop recruiting new participants for that reason.

**Figure 7.**
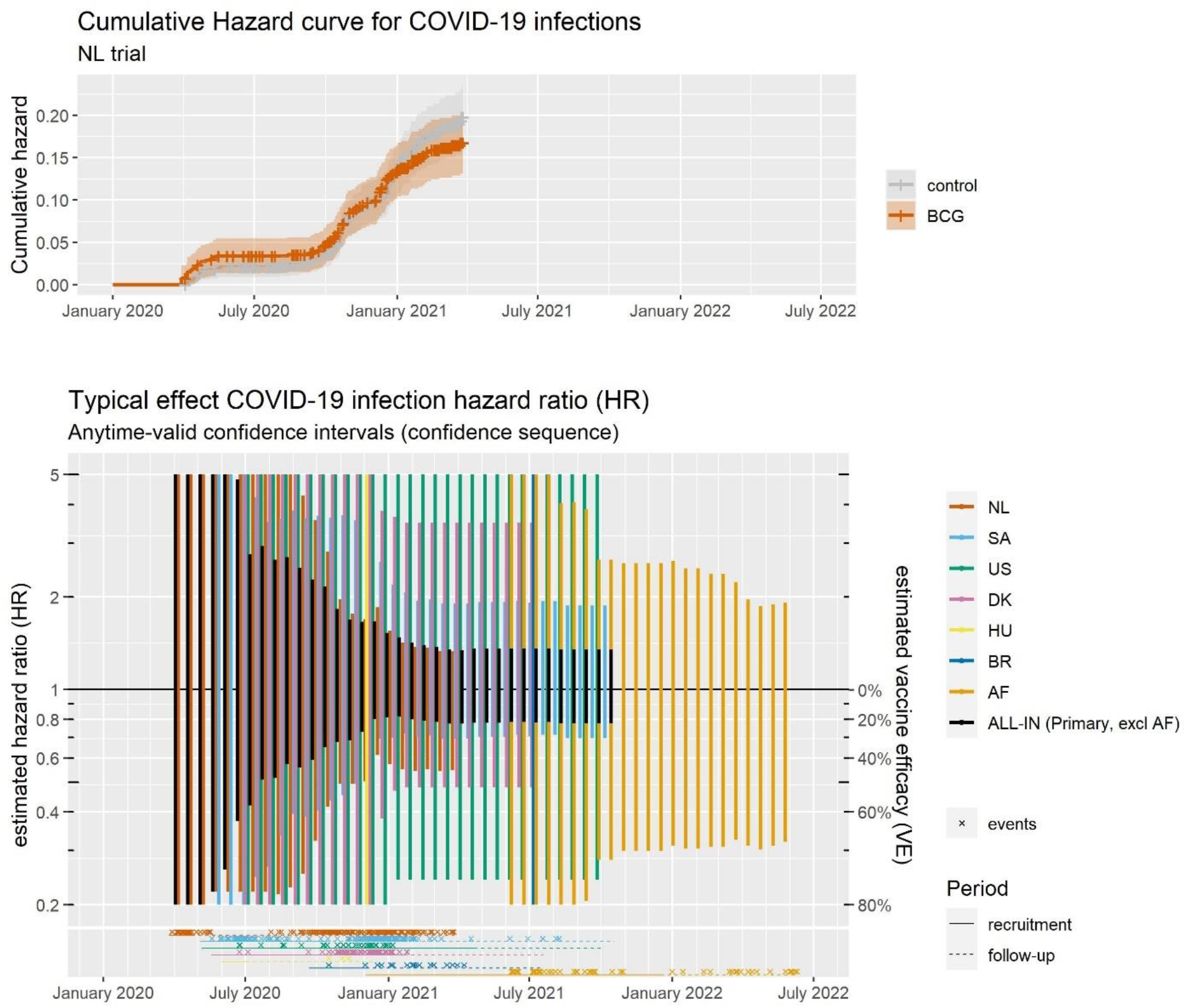
Sequences of anytime-valid confidence intervals for the trials and the meta-analysis from the first date with observed events of COVID-19 infection until the last date of follow-up. There is a confidence interval for every calendar day but these are only shown every 16 days for visibility. For HU and BR, intervals are only visible for the final day of follow-up, since these intervals stay the full plot width over time (larger than (0.2, 5), see Figure 5). AF observes most events at the 6-month end of follow-up serology for each participant. An example cumulative hazard plot is given for the NL trial to show how the incidence of COVID-19 infections in the two groups over time, and the censoring (indicated by +’es in the curve) relates to the HR estimates. The Dutch intervals can be seen in the background, shrinking fast between September 2020 and January 2021. Note that the y-axis is logarithmic.

A limitation of our study is that we were not able to include all trials and some trials were added only after they were concluded. This was mostly due to issues with the transfer of data, either from a legal or funder’s perspective. As a result, the meta-analysis had little chance to affect trial decisions. The use of aggregate data (recently proposed as an approach to prospective collaborative meta-analysis (Tierney, et al., 2021)) instead of individual participant data might have allowed a more timely and complete meta-analysis that could have had more impact during the pandemic. However, this would require each trial to generate and share these statistics and limits the data quality verification possibilities for the meta-analysis statistician. Therefore, it remains difficult to completely recommend against the IPD approach (Ter Schure, Grünwald, & Ly, 2021). One might add that, fortunately, specific COVID-19 vaccinations became available relatively quickly, in what was still an early stage of the ongoing meta-analysis. Had this not been the case, our meta-analysis would have been all the more urgent and data-transfer issues might have been overcome more easily. Since this is a live meta-analysis, efforts will be undertaken to keep this paper up to date as additional trials become available.

To the best of our knowledge, this is the first time a live meta-analysis of ongoing trials is being conducted on a continuous basis. A challenge when analysing data of ongoing trials is that datasets are subject to retrospective changes due to misclassifications and delayed registration of events. Considering misclassifications, we used objective outcome definitions to reduce this risk. Delayed registration cannot be avoided. Both misclassification and delayed registration could be argued to be conservative or result in unbiased relative effects if not associated with the intervention, but they may somewhat increase the type*-*I error rate. Despite these limitations, we are convinced that a live meta-analysis in an emergency setting with over 20 trials ongoing in parallel offers important potential benefits for society. Putative false*-*positive findings from one of these trials might have resulted in discontinuation of some of the other trials if they observed a trend in the same direction in their own data. Data from the meta-analysis would have protected trials against incorrect decisions in such circumstances.

Future studies should aim for a better understanding of how BCG-mediated changes in the immune system differentially affect SARS-CoV-2 and other respiratory infections. The role of BCG or other live vaccines in the immunogenicity of SARS-CoV-2 specific vaccines is also a topic of further research. From a methodological perspective, the continued development of anytime-valid meta-analysis techniques are likely to be extremely valuable for increasing the efficiency of global research efforts and reducing avoidable research waste (Ter Schure & Grünwald, 2022).

In conclusion, BCG vaccination has very little to no impact as an intervention for prevention of COVID-19 infections in healthcare workers, though the observation of very low numbers of severe cases (hospitalisations) prevented this study from measuring whether BCG vaccination has any impact on disease severity. Therefore, BCG should not be recommended as preventive intervention against COVID-19 infections in this population.

## Supporting information

PRISMA-IPD checklist

## Data Availability

All supplementary material and possibly links to data publications will be collected in this Replication Package:
https://www.researchequals.com/collections/kyep-h9

## APPENDIX

### Detailed methods

#### Anytime-validity without specifying a stopping rule

In contrast to methods based on *p*-values and conventional confidence intervals, analyses based on *e*-values and anytime-valid confidence intervals can be continuously monitored. Doing so would otherwise inflate type-I error rates for *p*-values (Armitage, McPherson, & Rowe, 1969), introduce accumulation bias in estimates for hazard ratios (Ter Schure & Grünwald, 2019), and lose coverage in conventional confidence intervals (Howard, Ramdas, McAuliffe, & Sekhon, 2021). With *e*-value methods, however, type-I error rates and coverage are preserved and any sequential or interim analysis can be validly performed, and trials can be added, without specifying an *alpha*-spending function or a maximum sample size (Ter Schure, Pérez-Ortiz, Ly, & Grünwald, 2022).

#### Type-I error control

Suppose that BCG is truly ineffective in preventing infections. Then, if we sample a sequence of events and record the corresponding logrank *e*-value for benefit after each event, the probability that this *e*-value sequence will *ever* increase above the threshold of 1/*α* = 400, is less than *α* = 0.025%, controlling the type-I error rate (chance of false-positives) at *α* = 0.025%, no matter how many new events are added to the analysis. Equivalently, suppose we independently sample (simulate) very many such sequences. In this ‘infinite repeated use’ less than *α* = 0.025% of the logrank *e-*value sequences for benefit will *ever* increase above the threshold of 1/*α* = 400. Similarly, if BCG is truly ineffective in preventing hospitalisations, in infinite repeated use less than *α* = 0.225% of logrank *e-*value sequences for benefit will *ever* increase above 1/*α* = 44 (Grünwald, De Heide, & Koolen, 2022; Vovk & Wang, 2021). This property allows us to freely monitor the *e-*value (updated whenever new data arrive) and stop for benefit as soon as the *e-*value crosses the threshold of 400 and 44, respectively, while retaining type-I error control.

#### Type-II error control

If BCG truly reduces COVID-19 infections or hospitalisations, then we would like to detect such an effect as quickly as possible. The fastest detecting *e-*value logrank test is known as the GROW *e-*value logrank test, and is tuned to a minimal clinically relevant (relative) reduction of COVID-19 risk. This design ensures that the *e*-values will grow based on data from any trial that observes that effect of minimal relevance, or an effect that is more extreme.

An *e*-value analysis can reach it threshold at any sample size, such that the ability of an *e*-value study design cannot be summarised by a single sample size and power. Each design has a dual sample size calculation: a maximum sample size and an average sample size that accompany a single power – for example 80%. The maximum sample size of an *e*-value logrank test is the number of events at which, under the minimal clinically relevant effect size, we have reached the threshold with 80% probability. Equivalently, we sample (simulate) a very large number of independent sequences of events with this effect size. Then the maximum sample size is the sample size at which, in this ’infinite repeated use’, 80% of the corresponding *e*-value sequences has reached a threshold. The average sample size is the average at which those *e-*value sequences reach that threshold, and is always smaller since many reach the threshold before the maximum sample size.

Such a dual sample size calculation shows that we need a maximum of 1345 events of COVID-19 infections to have 80% power for the *e-*value logrank test to reach the threshold of 400 for a minimal effect of HR 0.8 (simulated using designSafeLogrank(hrMin = 0.8, beta = 0.2, alpha= 0.0025, alternative = “less”), +-2x bootstrap se of 67) and will reach that threshold at 874 events on average (2x bootstrap se of 22)^1^. A similar dual sample size calculation shows that we need a maximum of approximately 355 events of COVID-19 hospitalisation to have 80% power for the *e-*value logrank test to reach the threshold of 44 (simulated using designSafeLogrank(hrMin = 0.7, beta = 0.2, alpha = 0.0225, alternative = “less”), +-2x bootstrap se of 20) and will reach that threshold at 212 events on average (2x bootstrap se of 6). Note that we are more likely to observe higher *e-*values (like in *Figure 1* on page 3, simulated with HR = 0.7) if the effects are more extreme then these minimum effects (0.7 is further away from 1 than hrMin = 0.8 for infections). Also, a standard logrank test would allow only a single analysis and needs 1069 events to detect a HR of 0.8 and 225 events to detect a HR of 0.7, which is less than the maximum sample size for the *e-*value logrank test, but more than the average sample size.

#### Code used for the analysis

The event times are considered in calendar time such that all participants within the same hospital at a given date are at risk of any events observed, regardless of their own time since randomisation (in R processed using the Surv() function from the survival package with type = “counting”). Late entries are left-truncated and participants that are lost to follow-up or vaccinated with a COVID-19 specific vaccine are right-censored. Data was analysed for *e-*values using the R functions designSafeLogrank() and safeLogrankTest() from the R package safestats, maximum-likelihood estimators for the HR were obtained using coxph() from the R package survival, and anytime-valid confidence sequences were obtained using inverse*-*variance weighted z-scores for log(HR) using computeConfidenceIntervalZ() from the R package safestats (Turner, Ly, Pérez-Ortiz, ter Schure, & and Grünwald, 2022).

### Detailed results

Table 13 shows that the exact *e-*values and the Gaussian approximation to the logrank statistics are very similar such that the *e-*values can be easily recalculated based on the logrank Z-score (Ter Schure, Pérez-Ortiz, Ly, & Grünwald, 2022). The Gaussian *e-*values are likelihood ratios of Gaussians comparing the HR of minimal clinical relevance to the null, e.g. the *e-*value for COVID-19 infections in trial NL can be recalculated as

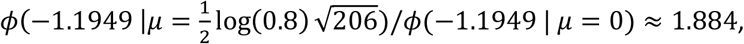

meaning that the data is hardly better supported by the alternative hypothesis of HR of 0.8 or smaller than by the null of HR 1.

**Table 13.**
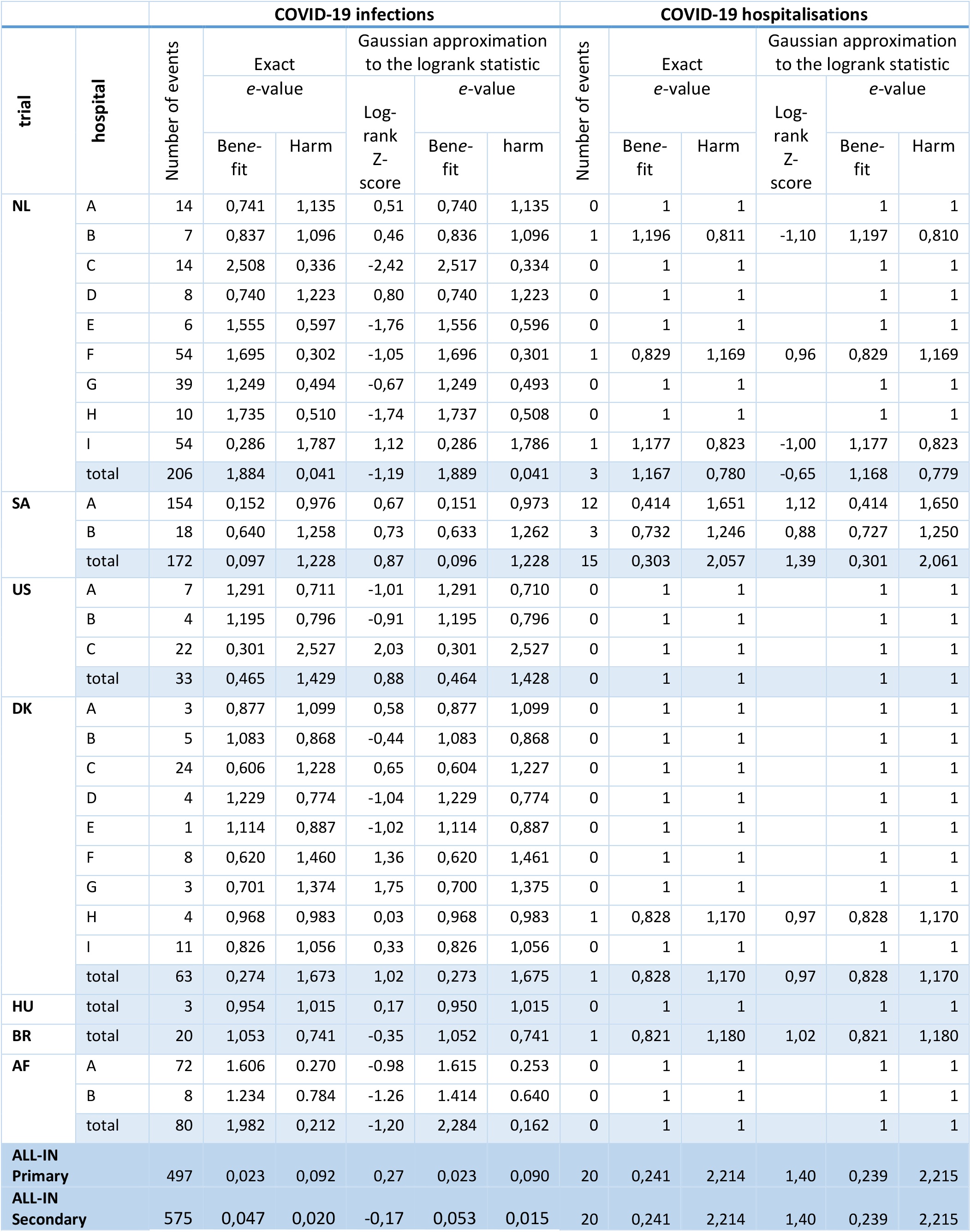
Exact and Gaussian logrank e-values by hospital within trial

|  |  | COVID-19 infections |  |  |  |  |  | COVID-19 hospitalisations |  |  |  |  |  |
| --- | --- | --- | --- | --- | --- | --- | --- | --- | --- | --- | --- | --- | --- |
| trial | hospital | Number of events | Exact |  | Gaussian approximation to the logrank statistic |  |  | Number of events | Exact |  | Gaussian approximation to the logrank statistic |  |  |
|  |  |  | e-value |  | Log-rank Z-score | e-value |  |  | e-value |  | Log-rank Z-score | e-value |  |
|  |  |  | Bene-fit | Harm |  | Bene-fit | harm |  | Bene-fit | Harm |  | Bene-fit | Harm |
| NL | A | 14 | 0,741 | 1,135 | 0,51 | 0,740 | 1,135 | 0 | 1 | 1 |  | 1 | 1 |
|  | B | 7 | 0,837 | 1,096 | 0,46 | 0,836 | 1,096 | 1 | 1,196 | 0,811 | -1,10 | 1,197 | 0,810 |
|  | C | 14 | 2,508 | 0,336 | -2,42 | 2,517 | 0,334 | 0 | 1 | 1 |  | 1 | 1 |
|  | D | 8 | 0,740 | 1,223 | 0,80 | 0,740 | 1,223 | 0 | 1 | 1 |  | 1 | 1 |
|  | E | 6 | 1,555 | 0,597 | -1,76 | 1,556 | 0,596 | 0 | 1 | 1 |  | 1 | 1 |
|  | F | 54 | 1,695 | 0,302 | -1,05 | 1,696 | 0,301 | 1 | 0,829 | 1,169 | 0,96 | 0,829 | 1,169 |
|  | G | 39 | 1,249 | 0,494 | -0,67 | 1,249 | 0,493 | 0 | 1 | 1 |  | 1 | 1 |
|  | H | 10 | 1,735 | 0,510 | -1,74 | 1,737 | 0,508 | 0 | 1 | 1 |  | 1 | 1 |
|  | I | 54 | 0,286 | 1,787 | 1,12 | 0,286 | 1,786 | 1 | 1,177 | 0,823 | -1,00 | 1,177 | 0,823 |
|  | total | 206 | 1,884 | 0,041 | -1,19 | 1,889 | 0,041 | 3 | 1,167 | 0,780 | -0,65 | 1,168 | 0,779 |
| SA | A | 154 | 0,152 | 0,976 | 0,67 | 0,151 | 0,973 | 12 | 0,414 | 1,651 | 1,12 | 0,414 | 1,650 |
|  | B | 18 | 0,640 | 1,258 | 0,73 | 0,633 | 1,262 | 3 | 0,732 | 1,246 | 0,88 | 0,727 | 1,250 |
|  | total | 172 | 0,097 | 1,228 | 0,87 | 0,096 | 1,228 | 15 | 0,303 | 2,057 | 1,39 | 0,301 | 2,061 |
| US | A | 7 | 1,291 | 0,711 | -1,01 | 1,291 | 0,710 | 0 | 1 | 1 |  | 1 | 1 |
|  | B | 4 | 1,195 | 0,796 | -0,91 | 1,195 | 0,796 | 0 | 1 | 1 |  | 1 | 1 |
|  | C | 22 | 0,301 | 2,527 | 2,03 | 0,301 | 2,527 | 0 | 1 | 1 |  | 1 | 1 |
|  | total | 33 | 0,465 | 1,429 | 0,88 | 0,464 | 1,428 | 0 | 1 | 1 |  | 1 | 1 |
| DK | A | 3 | 0,877 | 1,099 | 0,58 | 0,877 | 1,099 | 0 | 1 | 1 |  | 1 | 1 |
|  | B | 5 | 1,083 | 0,868 | -0,44 | 1,083 | 0,868 | 0 | 1 | 1 |  | 1 | 1 |
|  | C | 24 | 0,606 | 1,228 | 0,65 | 0,604 | 1,227 | 0 | 1 | 1 |  | 1 | 1 |
|  | D | 4 | 1,229 | 0,774 | -1,04 | 1,229 | 0,774 | 0 | 1 | 1 |  | 1 | 1 |
|  | E | 1 | 1,114 | 0,887 | -1,02 | 1,114 | 0,887 | 0 | 1 | 1 |  | 1 | 1 |
|  | F | 8 | 0,620 | 1,460 | 1,36 | 0,620 | 1,461 | 0 | 1 | 1 |  | 1 | 1 |
|  | G | 3 | 0,701 | 1,374 | 1,75 | 0,700 | 1,375 | 0 | 1 | 1 |  | 1 | 1 |
|  | H | 4 | 0,968 | 0,983 | 0,03 | 0,968 | 0,983 | 1 | 0,828 | 1,170 | 0,97 | 0,828 | 1,170 |
|  | I | 11 | 0,826 | 1,056 | 0,33 | 0,826 | 1,056 | 0 | 1 | 1 |  | 1 | 1 |
|  | total | 63 | 0,274 | 1,673 | 1,02 | 0,273 | 1,675 | 1 | 0,828 | 1,170 | 0,97 | 0,828 | 1,170 |
| HU | total | 3 | 0,954 | 1,015 | 0,17 | 0,950 | 1,015 | 0 | 1 | 1 |  | 1 | 1 |
| BR | total | 20 | 1,053 | 0,741 | -0,35 | 1,052 | 0,741 | 1 | 0,821 | 1,180 | 1,02 | 0,821 | 1,180 |
| AF | A | 72 | 1.606 | 0.270 | -0.98 | 1.615 | 0.253 | 0 | 1 | 1 |  | 1 | 1 |
|  | B | 8 | 1.234 | 0.784 | -1.26 | 1.414 | 0.640 | 0 | 1 | 1 |  | 1 | 1 |
|  | total | 80 | 1,982 | 0,212 | -1,20 | 2,284 | 0,162 | 0 | 1 | 1 |  | 1 | 1 |
| ALL-IN Primary |  | 497 | 0,023 | 0,092 | 0,27 | 0,023 | 0,090 | 20 | 0,241 | 2,214 | 1,40 | 0,239 | 2,215 |
| ALL-IN Secondary |  | 575 | 0,047 | 0,020 | -0,17 | 0,053 | 0,015 | 20 | 0,241 | 2,214 | 1,40 | 0,239 | 2,215 |

The *e*-values for the secondary analysis can be easily obtained by multiplying the primary analysis *e*-value of 0.023 by the AF *e*-value of 1.203 into 0.028.

The typical effect size for the HR is an inverse variance weighted estimate, e.g. (from *Table 14*) 1.02 is estimated as exp(0.02) following: 0.02 =

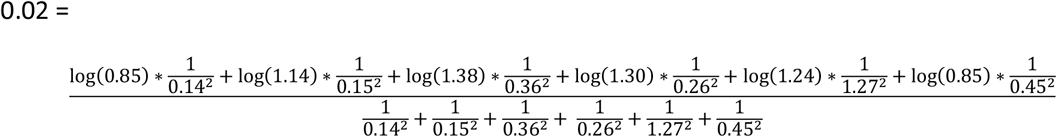

**Table 14.** Detailed estimates for COVID-19 infections

| COVID-19 infections |  |  |  |  |  |  |  |  |
| --- | --- | --- | --- | --- | --- | --- | --- | --- |
| Trial | Summary statistics |  |  | Hazard Ratio (HR) |  |  |  |  |
|  |  |  |  | Maximum likelihood estimator Cox model stratified by hospital Typical effect size | Anytime-valid 95% CI |  | Anytime-valid 99.5% CI |  |
|  | Number of events | Sum(Observed - Expected) | Standard error log(HR) |  | lower | upper | lower | upper |
| NL | 206 | -8,56 | 0,14 | 0,85 | 0,54 | 1,32 | 0,48 | 1,49 |
| SA | 172 | 5,69 | 0,15 | 1,14 | 0,70 | 1,87 | 0,60 | 2,16 |
| US | 31 | 2,50 | 0,36 | 1,38 | 0,24 | 7,97 | 0,14 | 13,84 |
| DK | 63 | 4,05 | 0,26 | 1,30 | 0,48 | 3,50 | 0,36 | 4,73 |
| HU | 3 | 0,14 | 1,27 | 1,24 | <0,01 | >99 | <0,01 | >99 |
| BR | 20 | -0,78 | 0,45 | 0,85 | 0,07 | 10,61 | 0,03 | 23,75 |
| AF | 80 | -5,04 | 0,23 | 0,76 | 0,32 | 1,81 | 0,25 | 2,35 |
| ALL-IN Pri-<br>mary | 495 | 3,03 | 0,09 | 1,02 | 0,78 | 1,35 | 0,73 | 1,45 |
| Sec-<br>ondary | 575 | -2,01 | 0,08 | 0,98 | 0,76 | 1,27 | 0,71 | 1,36 |

These estimates can also be approximated by the Peto estimator (Yusuf, Peto, Lewis, Collins, & Sleight, 1985, pp. 366-367, Statistical Appendix) based on the sum of observed minus expected (Sum(Observed - Expected)) and the approximate sum of variances of these observed minus expected, of n*(½)*(1 − ½),

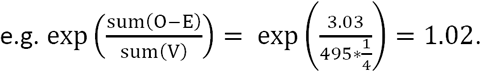

Statistical Appendix

### Anytime-valid confidence intervals for location parameters based on *Z*-scores

In this appendix we elaborate on the anytime-valid confidence intervals for a univariate parameter *θ* based on *Z*-scores as implemented in the safestats package (Turner, Ly, Ortiz-Perez, ter Schure, & Grünwald, 2022) and shown in the main text. We assume availability of an asymptotically normal estimator 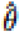 for scalar parameter *θ*. where we denote the value 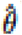 takes on a sample *Y*_1_,…,*Y*_2_ of length *n* by 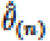 and its standard error by SE_(*N*)_. Thu (approximate or exact, see below) anytime-valid (1 - *α*)-confidence interval for *θ* that we employ is given by

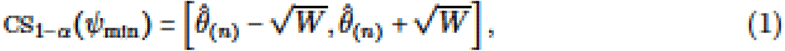

where we set

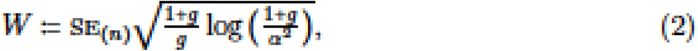

where 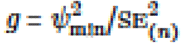 and *ψ*_min_ is a pre-set minimal clinically relevant (mean) difference parameter such as (as in the main text) *ψ*_min_ = log(0.8) _PD_ -0.223.

If the data *Y*_1_,*Y*_2_,… are i.i.d. normally distributed with fixed variance and mean *θ* and 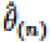 is the maximum likelihood estimator (MLE), then the confidence interval Eq. (1) is exact and exactly anytime-valid, a notion explained below. This holds for any fixed value of meta-parameter *ψ*_min_ picking *ψ*_min_ to be equal to a pre-specified minimum clinically relevant effect size merely serves to ‘fine-tune’ the intervals so that they are optimized to exclude any pre-specified null hypothesis early on (i.e. at small sample size) if the effect is at least as far away from that null hypothesis (in terms of mean difference) as |ψ_min_| 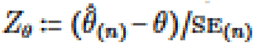 is well-approximated by a standard normal, then the interval is still approximately anytime-valid. This is the the case if *θ* represents the logarithm of the hazard ratio as in the main text and 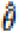 is either the MLE based on Cox’ partial likelihood with a single covariate treatment/control or, as in the main text, the fixed-effects inverse variance weighted effect size estimator typically used in meta-analysis.

#### Anytime-Valid Bayes Factors for the Normal Location Family

To derive the intervals above and get an idea of their width, we consider the exact case where the data come from the normal location family. Thus, let 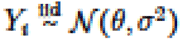 with *σ* known and *θ* the freely varying location (i.e.. mean). Let *θ*_0_ be a postulated value for the true value of the mean *θ*. Each such *θ*_0_ fixes the population mean at, for instance, *θ*_0_, = 0 or *θ*_0_, = −3.1415, and, for data *Y*_*1*_,…,*Y*_*n*_, yields a *relative* (to *θ*_0_,) *z*-statistic defined by 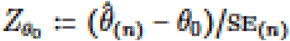 where 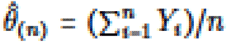 is the sample mean. and 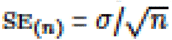 is the standard error.

For normal location problems Grünwald, de Heide, and Koolen (2019) showed that any Bayes factor for testing a simple null hypothesis provides an e-value. when evaluated at a fixed sample size *n*. and more generally, provides an *e-process* (Ramdas, Grünwald, Vovk, & Shafer, 2022) when evaluated at an arbitrary stopping time, leading, as we explain below, to anytime-valid tests. This holds in particular for Bayes factors in which the null represents *θ* = *θ*_0_ and the alternative is formed by a Bayesian prior; in particular we may take any conjugate, i.e. easily computable, prior of the form 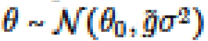 for any fixed tuning parameter 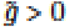. The resulting Bayes factor can then, by standard calculations (analogous to these of Example 3 of Grünwald et al.. 2019) be written, at sample size *n*. as

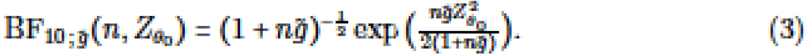

Anytime-validity relates to the behaviour of 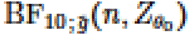 whenever the null hypothesis ℋ_*0*_ : *θ = θD* holds true. More specifically. the fact that 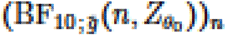 constitutes an e-proccss implies that, for any 0 < *α* < 1 (e.g. *α* = 0.05), the test that rejects ℋ_0_ whenever 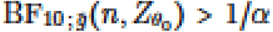, when *α* = 0.05) has a typo I error guarantee at level *α* irrespective of *n* being fixed in advanced, or informed by the observations. Equivalently, provided that *θ* is truly *θ*_*0*_ the statistic 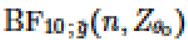 remains smaller than l/*θ* (i.e. smaller than 20 if *α* = 0.05) forever, with a least 1 − *α* (i.e. 05%. if *α* = 0.05) chance. This property holds irrespective of our choice of prior for the alternative, in particular for any fixed choice 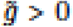 in (3): although we use Bayesian tools to derive our confidence intervals, their validity holds in a frequentist, non-Bayesian way.

Let us now fix 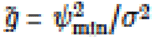 minimum relevant effect size *ψ*_min_ (we explain this choice for 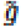 later). We now construct, for fixed *α*, the corresponding 1 − *α*-confidence interval CS_1−*α*(*n*)_(*ψ*_min_) at sample size *n* to consist of any *θ*_*0*_ such that 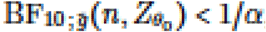, i.e. all *θ*_*0*_ that would not have been rejected by the test described above, had they been the null hypothesis. A straightforward computation shows that, with 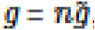, this gives the interval Eq. (1).

Any time-validity now moans the following: with probability at least 1−*α*, the true parameter value *θ* will be simultaneously contained in all intervals CS_1−*α*(*n*)_(*ψ*_min_), i.e. for *n* = 1. *n* = 2, *n* = 3 and so on. In particular, this means that this sequence of confidence intervals (or ^‘^confidence sequence’ as it is usually called, e.g. Howard, Ramdas, MeAuliffe. & Sekhon. 2021) will cover the true parameter value *θ* with at least (1 − *α*)% chance regardless of the specific-time (i.e. sample size *n*) we look at the data; we may also peek at the data as often as we like. Equivalently, we may imagine sampling (simulating) many independent sequences **Y**_1_ = *Y*_1,1_,*Y*_1,2_…,**Y**_**2**_ = ***Y***_**2**,**1**,_*Y*_2,2_,…, **Y**_**3**_ = ***Y***_**3**,**1**_,***Y***_**3**,**2**_,… and so on, and make a sequence of confidence intervals as above for each of these sequences. If we do this very many times, then in such an ^‘^infinite repeated use^’^, at least (1 − *α*)% of these interval sequences will cover the true parameter value *θ* at all sample sizes *n*. As such, we can monitor and act on an anytime-valid confidence interval at any moment in time without over-inflating the risk of detecting a false positive result due to repeated testing across time.

To explain our choice 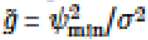, fix some *θ*_*0*_ representing the null in the test that we aim to accompany with an anytime-valid confidence interval (in the main text, with *θ*_*0*_ representing the log of the hazard ratio, this was just *θ*= 0 = log 1). We would like. among all anytime-valid confidence intervals constructed from (3) as above, the instance (i.e. the value of 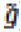) for which we ran expect the corresponding anytime-valid confidence interval to exclude *θ*_0_ as fast is possible (i.e. based on a sample that is as small as possible), whenever |*θ* − *θ*_0_| ≥ *ψ*_min_|. According to the theory of GRD e-variables and any time-valid tests as developed by Grünwald et al. (2015). this is given by the value of 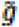 maximizing

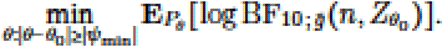

A straightforward computation shows that the maximum is achieved for 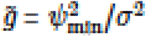 that 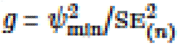 which is consistent with (1) above.

As stressed in the literature (Howard et al.. 2021), there is no such thing as an ‘overall optimal^’^ any time-valid confidence interval: the interval with the minimum widths at sample size n in a fixed sub-region of values for 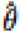 (say. if 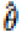 is close to *θ*_0_ + *ψ*_min_) will be suboptimal (overly wide) for other sample sizes and regions of 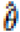 and may not even shrink to width zero as *n* increases. On the other hand, the one that shrinks fastest to 0 asymptotically may have overly large width at the smaller *n* of interest and cannot be easily calculated. By insisting on intervals generated by Bayes factors with mean centered at null, _*0*_ we obtain a compromise: easily calculable anytime-valid confidence intervals that have some sensitivity to minimal effect size *ψ*_min_ whereas at the same time the guarantee that the width at time *n* is of order 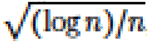, being wider than traditional fixed-*n* intervals by just a 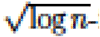 factor this guarantee follows by the same reasoning as in Example 3 of Grünwald et al. (2019)).

## Footnotes

1 These numbers (1 345 and 874) are much larger than the actual number (495) of COVID-19 infections we have observed so far, at the time of this publication, over all contributing trials. This might perhaps suggest that we should wait for hundreds of additional events and that our analysis, while valid at any number of events, is “not relevant yet”. Crucially though, it already *is* relevant: already at the current number of events we have a 95%-anytime-valid confidence interval 0.78-1.35, which means that we have almost reached the point where we can in principle stop, not for having reached sufficient power but instead for futility. As already stated in the main text, this happens as soon as the lower end of the confidence interval exceeds minimum relevant effect size 0.8 so that any effect size ≤ 0.8 is ruled out of our always-valid confidence interval (this effectively corresponds to all these hypotheses, rather than the null, being rejected).

