## Supplementary material for "Bacillus Calmette-Guérin vaccine to reduce COVID-19 infections and hospitalisations in healthcare workers – a living systematic review and prospective ALL-IN meta-analysis of individual participant data from randomised controlled trials": PRISMA-IPD checklist

### Supplementary Online Content

Stewart LA, Clarke M, Rovers M, et al; PRISMA-IPD Development Group. Preferred reporting items for a systematic review and meta-analysis of individual participant data: the PRISMA-IPD statement. *JAMA*. doi:10.1001/jama.2015.3656.

**eTable 1.** PRISMA-IPD Alignment With PRISMA

**eTable 2.** Examples of Reporting Systematic Reviews and Meta-analysis of IPD

**eReferences**

This supplementary material has been provided by the authors to give readers additional information about their work.

© 2015 American Medical Association. All rights reserved.

**eTable 1.** PRISMA-IPD alignment with PRISMA

| Section/ topic | Item | PRISMA Checklist item * | | PRISMA-IPD Checklist Item where different |
|---|---|---|---|---|
| Title | 1 | Identify the report as a systematic review, meta-analysis, or both. | | Identify the report as a systematic review and meta-analysis of individual participant data. |
| Structured summary ** | 2 | TITLE | 1. Title: Identify the report as a systematic review, meta-analysis, or both. | Provide a structured summary including as applicable:<br>**Background**: state research question and main objectives with information on participants, interventions, comparators and outcomes.<br>**Methods**: report eligibility criteria; data sources including dates of last bibliographic search or elicitation, noting that IPD were sought; methods of assessing risk of bias.<br>**Results**: provide number and type of studies and participants identified and number (%) obtained; summary effect estimates for main outcomes (benefits and harms) with confidence intervals and measures of statistical heterogeneity. Describe of the direction and size of summary effects in terms meaningful to those who would put findings into practice.<br>**Discussion:** state main strengths and limitations of the evidence, general interpretation of the results and any important implications.<br>**Other:** report primary funding source, registration number and registry name for the systematic review and IPD meta-analysis |
| | | BACKGROUND | 2. Objectives: The research question including components such as participants, interventions, comparators, and outcomes. | |
| | | METHODS | 3. Eligibility criteria: Study and report characteristics used as criteria for inclusion.<br>4. Information sources: Key databases searched and search dates.<br>5. Risk of bias: Methods of assessing risk of bias. | |
| | | RESULTS | 6. Included studies: Number and type of included studies and participants and relevant characteristics of studies.<br>7. Synthesis of results: Results for main outcomes (benefits and harms), preferably indicating the number of studies and participants for each. If meta-analysis was done, include summary measures and confidence intervals.<br>8. Description of the effect: Direction of the effect (i.e., which group is favored) and size of the effect in terms meaningful to clinicians and patients | |
| | | DISCUSSION | 9. Strengths and Limitations of evidence: Brief summary of strengths and limitations of evidence (e.g., inconsistency, imprecision, indirectness, or risk of bias, other supporting or conflicting evidence).<br>10. Interpretation: General interpretation of the results and important implications. | |
| Rationale | 3 | Describe the rationale for the review in the context of what is already known. | | |

© 2015 American Medical Association. All rights reserved.

| | | | |
|---|---|---|---|
| Objectives | 4 | Provide an explicit statement of questions being addressed with reference to participants, interventions, comparisons, outcomes, and study design (PICOS). | Provide an explicit statement of the questions being addressed with reference, as applicable, to participants, interventions, comparisons, outcomes and study design (PICOS). Include any hypotheses that relate to particular types of participant-level subgroups. |
| Protocol and registration | 5 | Indicate if a review protocol exists, if and where it can be accessed (e.g., Web address), and, if available, provide registration information including registration number. | Indicate if a protocol exists and where it can be accessed. If available, provide registration information including registration number and registry name. Provide publication details, if applicable. |
| Eligibility criteria | 6 | Specify study characteristics (e.g., PICOS, length of follow-up) and report characteristics (e.g., years considered, language, publication status) used as criteria for eligibility, giving rationale. | Specify inclusion and exclusion criteria including those relating to participants, interventions, comparisons, outcomes, study design and characteristics (e.g. years when conducted, required minimum follow-up). Note whether these were applied at the study or individual level i.e. whether eligible participants were included (and ineligible participants excluded) from a study that included a wider population than specified by the review inclusion criteria. The rationale for criteria should be stated. |
| Information sources<br>Identifying studies - information sources | 7 | Describe all information sources (e.g., databases with dates of coverage, contact with study authors to identify additional studies) in the search and date last searched. | Describe all methods of identifying published and unpublished studies including, as applicable: which bibliographic databases were searched with dates of coverage; details of any hand searching including of conference proceedings; use of study registers and agency or company databases; contact with the original research team and experts in the field; open advertisements and surveys. Give the date of last search or elicitation. |
| Search<br>Identifying studies - search | 8 | Present full electronic search strategy for at least one database, including any limits used, such that it could be repeated. | |
| Study selection | 9 | State the process for selecting studies (i.e., screening, eligibility, included in systematic review, and, if applicable, included in the meta-analysis). | State the process for determining which studies were eligible for inclusion. |
| Data collection process | 10 | Describe method of data extraction from reports (e.g., piloted forms, independently, in duplicate) and any processes for obtaining and confirming data from investigators. | Describe how IPD were requested, collected and managed, including any processes for querying and confirming data with investigators. If IPD were not sought from any eligible study, the reason for this should be stated (for each such study). |
| | | | If applicable, describe how any studies for which IPD were not available were dealt with. This should include whether, how and what aggregate data were sought or extracted from study reports and publications (such as extracting data independently in duplicate) and any processes for obtaining and confirming these data with investigators. |
| Data items | 11 | List and define all variables for which data were sought (e.g., PICOS, funding sources) and any assumptions and simplifications made. | Describe how the information and variables to be collected were chosen. List and define all study level and participant level data that were sought, including baseline and follow-up information. If applicable, describe methods of standardizing or translating variables within the IPD datasets to ensure common scales or |

© 2015 American Medical Association. All rights reserved.

| | | | |
|---|---|---|---|
| | | | measurements across studies. |
| A1<br>IPD Integrity | | | Describe what aspects of IPD were subject to data checking (such as sequence generation, data consistency and completeness, baseline imbalance) and how this was done. |
| Risk of bias in individual studies | 12 | Describe methods used for assessing risk of bias of individual studies (including specification of whether this was done at the study or outcome level), and how this information is to be used in any data synthesis. | Describe methods used to assess risk of bias in the individual studies and whether this was applied separately for each outcome. If applicable, describe how findings of IPD checking were used to inform the assessment. Report if and how risk of bias assessment was used in any data synthesis. |
| Summary measures<br>Specification of outcomes and effect measures | 13 | State the principal summary measures (e.g., risk ratio, difference in means). | State all treatment comparisons of interests. State all outcomes addressed and define them in detail. State whether they were pre-specified for the review and, if applicable, whether they were primary/main or secondary/additional outcomes. Give the principal measures of effect (such as risk ratio, hazard ratio, difference in means) used for each outcome. |
| Synthesis of results<br>Synthesis methods | 14 | Describe the methods of handling data and combining results of studies, if done, including measures of consistency (e.g., $I^2$) for each meta-analysis. | Describe the meta-analysis methods used to synthesize IPD. Specify any statistical methods and models used. Issues should include (but are not restricted to):<br>• Use of a one-stage or two-stage approach.<br>• How effect estimates were generated separately within each study and combined across studies (where applicable).<br>• Specification of one-stage models (where applicable), including how clustering of patients within studies was accounted for.<br>• Use of fixed or random effects models and any other model assumptions, such as proportional hazards.<br>• How (summary) survival curves were generated (where applicable).<br>• Methods for quantifying statistical heterogeneity (such as $I^2$ and $\tau^2$).<br>• How studies providing IPD and not providing IPD were analyzed together (where applicable).<br>• How missing data within the IPD were dealt with (where applicable). |
| A2<br>Exploration of variation in effects | | | If applicable, describe any methods used to explore variation in effects by study or participant level characteristics (such as estimation of interactions between effect and covariates) State all participant-level characteristics that were analyzed as potential effect modifiers, and whether these were pre-specified. |
| Risk of bias across studies | 15 | Specify any assessment of risk of bias that may affect the cumulative evidence (e.g., publication bias, selective reporting within studies). | Specify any assessment of risk of bias relating to the accumulated body of evidence, including any pertaining to not obtaining IPD for particular studies, outcomes or other variables. |
| Additional analyses | 16 | Describe methods of additional analyses (e.g., sensitivity or subgroup analyses, meta-regression), if done, indicating which were pre-specified. | Describe methods of any additional analyses, including sensitivity analyses. State which of these were pre-specified. |

© 2015 American Medical Association. All rights reserved.

| | | | |
|---|---|---|---|
| Study selection<br>Study selection and IPD obtained | 17 | Give numbers of studies screened, assessed for eligibility, and included in the review, with reasons for exclusions at each stage, ideally with a flow diagram. | Give numbers of studies screened, assessed for eligibility, and included in the systematic review with reasons for exclusions at each stage. Indicate the number of studies and participants for which IPD were sought and for which IPD were obtained. For those studies where IPD were not available, give the numbers of studies and participants for which aggregate data were available. Report reasons for non-availability of IPD. Include a flow diagram. |
| Study characteristics | 18 | For each study, present characteristics for which data were extracted (e.g., study size, PICOS, follow-up period) and provide the citations. | For each study, present information on key study and participant characteristics (such as description of interventions, numbers of participants, demographic data, unavailability of outcomes, funding source, and if applicable duration of follow-up). Provide (main) citations for each study. Where applicable, also report similar study characteristics for any studies not providing IPD. |
| A3<br>IPD integrity | | | Report any important issues identified in checking IPD or state that there were none. |
| Risk of bias within studies | 19 | Present data on risk of bias of each study and, if available, any outcome level assessment (see item 12). | Present data on risk of bias assessments. If applicable, describe whether data checking led to the up-weighting or down-weighting of these assessments. Consider how any potential bias affects the robustness of meta-analysis conclusions. |
| Results of individual studies | 20 | For all outcomes considered (benefits or harms), present, for each study: (a) simple summary data for each intervention group (b) effect estimates and confidence intervals, ideally with a forest plot. | For each comparison and for each main outcome (benefit or harm), for each individual study report the number of eligible participants for which data were obtained and show simple summary data for each intervention group (including, where applicable, the number of events), effect estimates and confidence intervals. These may be tabulated or included on a forest plot. |
| Synthesis of results<br>Results of syntheses | 21 | Present results of each meta-analysis done, including confidence intervals and measures of consistency. | Present summary effects for each meta-analysis undertaken, including confidence intervals and measures of statistical heterogeneity. State whether the analysis was pre-specified, report the numbers of studies and participants, and, where applicable, the number of events on which it is based. |
| | | | When exploring variation in effects due to patient or study characteristics, present summary interaction estimates for each characteristic examined, including confidence intervals and measures of statistical heterogeneity. State whether the analysis was pre-specified. State whether any interaction is consistent across trials. |
| | | | Provide a description of the direction and size of effect in terms meaningful to those who would put findings into practice. |
| Risk of bias across studies | 22 | Present results of any assessment of risk of bias across studies (see Item 15). | Present results of any assessment of risk of bias relating to the accumulated body of evidence, including any pertaining to the availability and representativeness of available studies, outcomes or other variables. |

© 2015 American Medical Association. All rights reserved.

| | | | |
|---|---|---|---|
| Additional analysis | 23 | Give results of additional analyses, if done (e.g., sensitivity or subgroup analyses, meta-regression [see Item 16]). | Give results of any additional analyses (e.g. sensitivity analyses). If applicable, this should also include any analyses that incorporate aggregate data for studies that do not have IPD. |
| | | | If applicable, summarize the main meta-analysis results following the inclusion or exclusion of studies for which IPD were not available. |
| Summary of evidence | 24 | Summarize the main findings including the strength of evidence for each main outcome; consider their relevance to key groups (e.g., healthcare providers, users, and policy makers). | Summarize the main findings, including the strength of evidence for each main outcome. |
| Limitations<br>Strengths and limitations | 25 | Discuss limitations at study and outcome level (e.g., risk of bias), and at review-level (e.g., incomplete retrieval of identified research, reporting bias). | Discuss any important strengths and limitations of the evidence. |
| Conclusions | 26 | Provide a general interpretation of the results in the context of other evidence, and implications for future research. | Provide a general interpretation of the findings in the context of other evidence. |
| A4<br>Implications | | | Consider relevance to key groups (such as policy makers, service providers and service users). Consider implications for future research. |
| Funding | 27 | Describe sources of funding for the systematic review and other support (e.g., supply of data); role of funders for the systematic review. | Describe sources of funding and other support (such as supply of IPD), and the role in the systematic review of those providing such support. |

eTable 1: PRISMA Checklist and changes for PRISMA-IPD **From:* Moher D, Liberati A, Tetzlaff J, Altman DG, The PRISMA Group (2009). Preferred Reporting Items for Systematic Reviews and Meta-Analyses: The PRISMA Statement. PLoS Med 6(6): e1000097. doi:10.1371/journal.pmed1000097 *** From:* Beller EM, Glasziou PP, Altman DG, Hopewell S, Bastian H, et al. (2013) PRISMA for Abstracts: Reporting Systematic Reviews in Journal and Conference Abstracts. PLoS Med 10(4): e1001419.

© 2015 American Medical Association. All rights reserved.

**eTable 2.** Examples of Reporting Systematic Reviews and Meta-analysis of IPD [a]

| Section/topic | Item | PRISMA-IPD Checklist item | Examples of reporting |
|---|---|---|---|
| **Title** | | | |
| Title | 1 | Identify the report as a systematic review and meta-analysis of individual participant data. | *From example study 1: cervical cancer*<br>Reducing uncertainties about the effects of chemoradiotherapy for cervical cancer: A systematic review and meta-analysis of individual patient data from 18 randomized trials. (Chemoradiotherapy for Cervical Cancer Meta-analysis Collaboration 2008) [1] |
| **Abstract** | | | |
| Structured summary | 2 | Provide a structured summary including as applicable: | *From example study 2: preoperative chemotherapy in non small cell lung cancer* |
| | | **Background**: state research question and main objectives with information on participants, interventions, comparators and outcomes. | **Background** Individual participant data meta-analyses of postoperative chemotherapy have shown improved survival for patients with non-small-cell lung cancer (NSCLC). We aimed to do a systematic review and individual participant data meta-analysis to establish the effect of preoperative chemotherapy for patients with resectable NSCLC. |
| | | **Methods**: report eligibility criteria; data sources including dates of last bibliographic search or elicitation, noting that IPD were sought; methods of assessing risk of bias. | **Methods** We systematically searched for trials that started after January, 1965. Updated individual participant data were centrally collected, checked, and analyzed. Results from individual randomized controlled trials (both published and unpublished) were combined using a two-stage fixed-effect model. Our primary outcome, overall survival, was defined as the time from randomization until death (any cause), with living patients censored on the date of last follow-up. Secondary outcomes were recurrence-free survival, time to locoregional and distant recurrence, cause-specific survival, complete and overall resection rates, and postoperative mortality. Prespecified analyses explored any variation in effect by trial and patient characteristics. All analyses were by intention to treat. |
| | | **Results**: provide number and type of studies and participants identified and number (%) obtained; summary effect estimates for main outcomes (benefits and harms) with confidence intervals and measures of statistical heterogeneity. Describe of the direction and size of summary effects in terms meaningful to those who would put findings into practice. | **Findings** Analyses of 15 randomized controlled trials (2385 patients) showed a significant benefit of preoperative chemotherapy on survival (hazard ratio [HR] 0·87, 95% CI 0·78–0·96, p=0·007), a 13% reduction in the relative risk of death (no evidence of a difference between trials; p=0·18, I2=25%). This finding represents an absolute survival improvement of 5% at 5 years, from 40% to 45%. There was no clear evidence of a difference in the effect on survival by chemotherapy regimen or scheduling, number of drugs, platinum agent used, or whether postoperative radiotherapy was given. There was no clear evidence that particular types of patient defined by age, sex, performance status, histology, or clinical stage benefited more or less from preoperative chemotherapy. Recurrence-free survival (HR 0·85, 95% CI 0·76–0·94, p=0·002) and time to distant recurrence (0·69, 0·58–0·82, p<0·0001) results were both significantly in favor of preoperative chemotherapy although most patients included were stage IB–IIIA. Results for time to locoregional recurrence (0·88, 0·73–1·07, p=0·20), although in favor of preoperative |

© 2015 American Medical Association. All rights reserved.

| | | | |
|---|---|---|---|
| | | | chemotherapy, were not statistically significant. Findings, which are based on 92% of all patients who were randomized, and mainly stage IB–IIIA, show preoperative chemotherapy significantly improves overall survival, time to distant recurrence, and recurrence- free survival in resectable NSCLC. The findings suggest this is a valid treatment option for most of these patients. Toxic effects could not be assessed. |
| | | **Discussion:** state main strengths and limitations of the evidence, general interpretation of the results and any important implications. | |
| | | **Other:** report primary funding source, registration number and registry name for the systematic review and IPD meta-analysis. | **Funding** Medical Research Council UK. (NSCLC Collaborative Group 2014) [2] |
| **Introduction** | | | |
| Rationale | 3 | Describe the rationale for the systematic review in the context of what is already known | *From example study 3: advanced bladder cancer*<br><br>A previous systematic review of published trials concluded that there was no good evidence to suggest that adjuvant chemotherapy improved the survival of patients with invasive bladder cancer. […] A subsequent review of four trials that used cisplatin-based combination chemotherapy concluded that the trials provided insufficient evidence to support the routine use of this type of adjuvant chemotherapy in invasive bladder cancer. Criticisms raised by these reviewers related to the design, analysis and reporting of the trials. Firstly, all of the individual trials were underpowered to detect moderate differences between the two arms. […]Therefore in June 2001, we initiated a systematic review and meta-analysis of individual patient data (IPD) […] using this methodology, we aimed to provide a better evidence base with which to judge the effect of adjuvant chemotherapy on invasive bladder cancer. (Advanced Bladder Cancer Meta-analysis Collaboration 2005) [3] |
| Objectives | 4 | Provide an explicit statement of the questions being addressed with reference, as applicable, to participants, interventions, comparisons, outcomes and study design (PICOS). Include any hypotheses that relate to particular types of participant level (subgroups). | *From example study 4: radiotherapy in lung cancer*<br><br>The Meta-Analysis of Radiotherapy in Lung Cancer collaborative group decided to perform an individual patient data meta-analysis to accurately estimate the effect of modified RT on survival outcomes and toxicity and to distinguish between ineffective treatment and moderate treatment effects, which may be clinically relevant. (Mauguen A. et al. 2012) [4] |
| | | | *From example study 5: otitis media*<br><br>Independent predictors of an extended course of disease had been established in an earlier study within the same setting (unpublished data). We used these independent baseline predictors i.e. age (<2 *vs* ≥2 years), fever (yes *vs* no), and bilateral acute otitis media (yes *vs* no) to investigate whether those at risk of |

© 2015 American Medical Association. All rights reserved.

| | | | |
|---|---|---|---|
| | | | an extended course had enhanced benefits from treatment with antibiotics. We also examined the effects of concurrent otorrhoea at baseline (yes *vs* no), both alone and in combination with the identified predictors, since this condition seems to be a clinically relevant outcome that occurs too infrequently to be identified as an independent predictor. (Rovers et al. 2006) [5] |
| **Methods** | | | |
| Protocol and registration | 5 | Indicate if a protocol exists, if and where it can be accessed. If available, provide registration information including registration number and registry name. Provide publication details, if applicable. | *Example study 6: spinal fusion*<br><br>Methods were pre-specified in a protocol (supplement 1) that was registered in PROSPERO in February 2012 (CRD42012001907). (Simmonds et al. 2013) [6] |
| Eligibility criteria | 6 | Specify inclusion and exclusion criteria including those relating to participants, interventions, comparisons, outcomes, study design and characteristics (e.g. years when conducted, required minimum follow-up). Note whether these were applied at the study or individual level i.e. whether eligible participants were included (and ineligible participants excluded) from a study that included a wider population than specified by the review inclusion criteria. The rationale for criteria should be stated. | *From example study 4: radiotherapy in lung cancer*<br><br>To be eligible, trials were to include patients with non-metastatic lung cancer, randomly assigned in a way that precluded prior knowledge of treatment assignment. They had to compare modified radiotherapy (accelerated, hyperfractionated, or both) with conventional radiotherapy (five daily 1.8- to 2-Gy fractions per week and a minimum total dose of 40 Gy for SCLC and 60 Gy ) for NSCLC. Trials must have accrued between January 1, 1970, and December 31, 2005, and not be confounded by additional therapeutic differences between the two arms. Trials combining chemotherapy (CT) with radiotherapy were included only if CT doses and schedule were the same in the two arms. (Mauguen A. et al. 2012). [4] |
| Identifying studies - information sources | 7 | Describe all methods of identifying published and unpublished studies including, as applicable: which bibliographic databases were searched with dates of coverage, details of any hand searching including conference proceedings, use of study registers and agency or company databases, contact with the original research team and experts in the field, open advertisements and surveys. Give the date of last search or elicitation. | *Example study 6: spinal fusion*<br><br>We performed a systematic literature search of the Cochrane Central Register of Controlled Trials, MEDLINE, EMBASE, and Science Citation Index in January 2012 and automated "current awareness" searches up to June 2012.We also searched Clinical Trials.gov to identify ongoing or unpublished randomized trials and published a call for evidence. (Simmonds et al. 2013). [6] |
| | | | *From example study 1: cervical cancer*<br><br>To avoid publication bias, published and unpublished trials were included in the meta-analysis. We searched MEDLINE and CancerLit using an optimal search strategy, and also LILACS, the Physicians' Data Query, and other trials registers. These were supplemented from reference lists of identified trial reports and review articles and from meeting proceedings (International Gynecologic Cancer Society and the Society for Gynecologic Oncology, 1994 through 2007). Furthermore, all participating investigators were |

© 2015 American Medical Association. All rights reserved.

| | | | |
|---|---|---|---|
| | | | asked to supplement our provisional list of trials. Searches were regularly updated until November 2007. (Chemoradiotherapy for Cervical Cancer Meta-analysis Collaboration 2008). [1] |
| Identifying studies - Search | 8 | Present the full electronic search strategy for at least one database, including any limits used, such that it could be repeated. | Detailed search strategies are often provided in an appendix. |
| Study selection processes | 9 | State the process for determining which studies were eligible for inclusion. | *Example study 7: pre-eclampsia*<br><br>Each potentially eligible study was assessed independently by at least two members of the steering group, un-blinded to authorship. Any differences of opinion regarding the assessment of the inclusion criteria were resolved by discussion. (Askie et al. 2007). [7] |
| Data collection processes | 10 | Describe how IPD were requested, collected and managed, including any processes for querying and confirming data with investigators. If IPD were not sought from any eligible study, the reason for this should be stated (for each such study). | *Example study 7: pre-eclampsia*<br><br>Anonymized data for each of the pre-specified variables were requested for each woman randomized. Data were supplied in a variety of formats, re-coded as necessary, and were checked for internal consistency, consistency with published reports, and for missing items. Information about the trials—e.g. randomization method and antiplatelet dose were cross-checked with published reports, trial protocols, and data collection sheets. (Askie et al. 2007) [7] |
| | | If applicable, describe how any studies for which IPD were not available were dealt with. This should include whether, how and what aggregate data were sought or extracted from study reports and publications (such as extracting data independently in duplicate) and any processes for obtaining and confirming these data with investigators. | *Example study 8: non-Hodgkin Lymphoma*<br><br>If individual patient data (IPD) were not available, data were extracted from published results, e.g. survival curves, using methods described by {Parmar et al.} (Greb et al 2009). [8] |
| Data items | 11 | Describe how the information and variables to be collected were chosen. | *Example study 7: pre-eclampsia*<br><br>Data to be collected were agreed after extensive consultation within the PARIS Collaborative Group. (Askie et al 2007) [7] |
| | | List and define all study level and participant level data that were sought, including baseline and follow-up information. | Example study 9: multiple myeloma<br><br>The diagnostic data requested for each randomized patient were as follows: patient identifier, date of diagnosis, date of birth (or age at randomization), sex, Durie-Salmon stage, hemoglobin level, platelet count, WBC count, 32-microglobulin level, M-band type, creatinine concentration, calcium and albumin |

© 2015 American Medical Association. All rights reserved.

| | | | |
|---|---|---|---|
| | | | levels, presence of bone lesions, and performance status. The event data requested were as follows: date randomized, allocated treatment, type of response (complete, partial) and date of response, achievement and date of plateau phase, recurrence and its date, current status (alive, dead), and date of death or date of last follow-up evaluation. (The Myeloma Trialists' Collaborative Group 1998) [9] |
| | | | *Example study 10: non diabetic renal disease*<br><br>The "begin date" for each study was defined as the time at which treatment with study medications was begun. The "end date" was defined as the time at which the patient reached an outcome or completed the specified follow-up period. Values assigned for baseline blood pressure and serum creatinine concentration were measured within 3 months before the study begin date. Baseline urinary protein excretion was measured within 4 months before the begin date. The value nearest the begin date was considered the baseline value. Follow-up values for systolic and diastolic blood pressure, serum creatinine concentration, and urinary protein excretion were also recorded. Supine systolic and diastolic blood pressure were measured after 5 to 10 minutes of rest in all studies except one {14}, which provided only sitting blood pressure readings. Laboratory methods of measuring serum creatinine concentration and urinary protein excretion varied across studies. Two {18, 20} studies per- formed dipstick assessment of urinary protein and performed quantitative measurement only if the dipstick test result was positive. For these two studies, all "dipstick-negative" results were assigned a value of 0.1 g/d. In all other studies, urinary protein excretion of 0.1 g/d or less was assigned a value of 0.1 g/d. Values greater than 0.1 g/d were recorded as the exact values reported in the study.<br><br>Ethnicity was recorded for all patients but was not classified uniformly. For our analysis, ethnicity was classified as African American (black) or nonblack. Renal biopsy was not performed routinely. The cause of renal disease was determined by nephrologists at each clinical center on the basis of history, physical examination, urinalysis, and other laboratory tests and was classified as diabetic renal disease (type 2 diabetes mellitus), glomerular diseases, tubulointerstitial disease, polycystic kidney disease, hypertensive nephrosclerosis, and other causes of renal diseases. (Jafar et al. 2001). [10] |
| | | If applicable, describe methods of standardizing or translating variables within the IPD datasets to ensure common scales or measurements across studies. | *Example study 11: post operative radiotherapy in non small cell lung cancer*<br><br>Since stage was recorded by different classification systems in different trials, for the purposes of this meta-analysis, stage data were translated to a common staging system {table 1}. (Burdett and Stewart 2005). [11] |
| IPD integrity | A1 | Describe what aspects of IPD were subject to data checking (such as sequence generation, data consistency, baseline imbalance, and completeness) and how this was done. | *Example study 12: adjuvant chemotherapy for non small cell lung cancer*<br><br>We used standard checks to identify missing data, assess data validity, and consistency. We verified the amount of missing data, checked the order of dates, and assessed data validity and consistency. To assess randomization integrity, we checked patterns of treatment allocation and balance of baseline |

© 2015 American Medical Association. All rights reserved.

| | | | |
|---|---|---|---|
| | | | characteristics by treatment group. Follow-up of surviving patients was checked to ensure that it was balanced by treatment group and was up-to-date. Any queries were resolved and the final database verified by each trial investigator or statistician. (NSCLC Meta-analyses Collaborative Group 2010). [12] |
| Risk of bias assessment in individual studies. | 12 | Describe methods used to assess risk of bias in the individual studies and whether this was applied separately for each outcome. | *Example study 6: spinal fusion*<br>We assessed risk of bias by using the Cochrane Collaboration's "risk-of bias" tool in the RCTs and a modified form of the Newcastle-Ottawa Scale for nonrandomized studies {see Appendix 2 for details}. Risk of bias was assessed by at least 2 researchers independently, with disagreements resolved by discussion. (Simmonds et al. 2013). [6] |
| | | If applicable, describe how findings of IPD checking were used to inform the assessment. Report if and how risk of bias assessment was used in any data synthesis. | *From example study 2: preoperative chemotherapy in non small cell lung cancer*<br>Patterns of treatment allocation and the balance of baseline characteristics by treatment group were used to check randomization integrity and follow-up of surviving patients was checked to ensure it was up-to-date and balanced by arm and fed into a risk of bias assessment for each trial. (NSCLC Meta-analysis Collaborative Group 2014). [2] |
| Specification of outcomes and effect measures | 13 | State all treatment comparisons of interests. State all outcomes addressed and define them in detail. State whether they were pre-specified for the review and, if applicable, whether they were primary/main or secondary/additional outcomes. Give the principal measures of effect (such as risk ratio, hazard ratio, difference in means) used for each outcome. | *From example study 1: cervical cancer*<br>The primary outcome, overall survival, was defined as the time from randomization until death by any cause. Living patients were censored on the date of last follow-up [… ]For survival and recurrence outcomes, individual times to event were used to obtain hazard ratio (HR) estimates of treatment effect for individual trials [...], For binary outcomes of response and toxicity, the number of events and numbers of patients were used to calculate Peto odds ratio estimates of treatment effect for individual trials, which were pooled across trials using the stratified-by-trial, fixed-effect model. (Chemoradiotherapy for Cervical Cancer Meta-analysis Collaboration.2008). [1] |
| | | | *Example study 13: cancer risk with tumor necrosis factor (TNF) inhibitors*<br>To identify all potential cancer events, irrespective of report to a regulatory agency or inclusion in the published report of the trial, each sponsor's clinical and safety database was searched using a list of over 1800 pre-defined terms. These terms included defined cancer types, terms for conditions potentially harboring an invasive cancer (e.g., 'gammopathy'), and terms not more specific than neoplasia-related text strings such as 'neopl'. The study period encompassed trial initiation until 30 days after the planned end of treatment. Person-time at risk was calculated from start of treatment to the event date, or (if no event) to the latest of date of withdrawal, date of last personal contact with the patient, or 30 days after end of planned treatment. For all events indicating a possible malignancy, sponsors submitted narratives using a pre-defined standardized format that described the history and course of the event. Narratives were subsequently blinded with respect to sponsor and treatment assignment. Based on the information in |

© 2015 American Medical Association. All rights reserved.

| | | | |
|---|---|---|---|
| | | | these narratives two oncologist adjudicators determined the probabilities (definite, probable, possible, unlikely) that the event was (i) a cancer including its site, (ii) prevalent at the trial start, and (iii) a recurrence of a pre-trial cancer. The earliest reported date of clinical evidence of the cancer and the date of clinical diagnosis were also abstracted. Each adjudicator reviewed all narratives. Consensus was required. Based on the above probabilities, outcome was examined using three nested cancer definitions {see Appendix}. Outcome A included all cancer events (definite or probable cancers) diagnosed during the study period, using the date of diagnosis as the event date. Outcome B excluded from A those events, in retrospect, judged definitely prevalent on the basis of a first reported date of sign or symptom pre-dating the trial start. Outcome C excluded from B those events which, for other reasons than first date of sign/ symptom, were judged by the oncologists to be probably prevalent at trial start. (Askling et al 2011). [13] |
| Synthesis methods | 14 | Describe the meta-analysis methods used to synthesize IPD. Specify any statistical methods and models used. Issues should include (but are not restricted to):<br>• Use of a one-stage or two-stage approach.<br>• How effect estimates were generated separately within each study and combined across studies (where applicable).<br>• Specification of one-stage models (where applicable), including how clustering of patients within studies was accounted for.<br>• Use of fixed or random effects models and any other model assumptions, such as proportional hazards.<br>• How (summary) survival curves were generated (where applicable).<br>• Methods for quantifying statistical heterogeneity (such as $I^2$ and $\tau^2$).<br>• How studies providing IPD and not providing IPD were analyzed together (where applicable).<br>• How missing data within the IPD were dealt with (where applicable). | *Example study 12: adjuvant chemotherapy for non small cell lung cancer*<br>Unless otherwise stated, all analyses were pre-specified in the protocols, and undertaken on an intention-to-treat basis. For every outcome, we used the log-rank expected number of events and variance to calculate individual trial HRs, which were pooled across trials with the fixed effect model. Survival is also presented with simple (non-stratified) Kaplan-Meier curves. We calculated absolute differences in overall survival at 5 years using overall HRs and survival in the control group. If a difference in effect by trial group or patient subgroup was identified, we used HRs and control group survival for the relevant groups to calculate absolute differences; otherwise the overall HR was used. (NSCLC Meta-analyses Collaborative Group 2010). [12]<br><br>*Example study 15: early breast cancer*<br>Survival curves show time to recurrence, breast cancer mortality, and any mortality. Yearly rates of breast cancer mortality assess the excess mortality when the mortality rate in women without recurrence is subtracted from the overall mortality rate in all women. Correspondingly, rate ratios (RRs) for breast cancer mortality are estimated from log-rank analyses of mortality with recurrence, obtained by subtraction of the log-rank analyses of mortality without recurrence (i.e., censored at recurrence) from those of all mortality. (Early Breast Cancer Trialists' Collaborative Group 2011). [14]<br><br>*Example study 6: spinal fusion*<br>{Appendix 2} provides details of statistical methods and analyses. In the analyses of effectiveness for continuously distributed outcomes (such as ODI score), we calculated mean differences between treatment groups in the change in score from preoperative values. For dichotomous outcomes (such as successful fusion), we calculated relative risks (RRs). Both were calculated separately for every trial at every time point. We then used standard random-effects meta-analytic techniques to combine effect estimates across trials. Separate meta-analyses were done for each of the specified time points. Linear and logistic random-effects regression models were used to combine all data from all trials in “1-stage” meta- |

© 2015 American Medical Association. All rights reserved.

| | | | |
|---|---|---|---|
| | | | analyses as sensitivity analyses. Heterogeneity was assessed in all meta-analyses by using the Higgins $I^2$ statistic and the Cochran Q test. (Simmonds et al. 2013). [6] |
| Exploration of variation in effects | A2 | If applicable, describe any methods used to explore variation in effects by study or participant level characteristics (such as estimation of interactions between effect and covariates) State all participant-level characteristics that were analyzed as potential effect modifiers, and whether these were pre-specified. | *Example study 12: adjuvant chemotherapy for non small cell lung cancer*<br>To investigate differences in the treatment effect across patient subgroups, we undertook Cox regressions including the relevant treatment by subgroup interaction term within trials and the interaction coefficients (HRs) pooled across trials. $\chi^2$ tests and the $I^2$ statistic were used to assess heterogeneity in the treatment effect or patient subgroup interactions across trials. (NSCLC Meta-analyses Collaborative Group 2010) [12] |
| | | | *Example study 6: spinal fusion*<br>We performed a subgroup analysis (stratified by trial) to examine whether effects varied according to the type of spinal surgery or by rhBMP-2 formulation (INFUSE or AMPLIFY). We investigated whether patient-level factors (age, sex, smoking, alcohol consumption, body mass index, diabetic status, and history of spinal surgery for back pain) were associated with the effectiveness of rhBMP-2 surgery by using a 1-stage random-effects regression model that included interaction terms between patient-level factors and treatment. (Simmonds et al. 2013). [6] |
| Risk of bias across studies | 15 | Specify any assessment of risk of bias relating to the accumulated body of evidence, including any pertaining to not obtaining IPD for particular studies, outcomes or other variables. | *From example study 1: cervical cancer*<br>Where IPD were not available, wherever possible, we calculated HRs and associated statistics from reported time-to-event analyses and considered the impact on the analyses of IPD. (Chemoradiotherapy for Cervical Cancer Meta-analysis Collaboration.2008). [1] |
| Additional analyses | 16 | Describe methods of any additional analyses including sensitivity analyses. State which of these were pre-specified. | *From example study 1: cervical cancer*<br>The main analyses described were limited to trials that compared concomitant chemotherapy and radical radiotherapy (with or without surgery) with the same radical radiotherapy (with or without surgery). However, to establish how sensitive the effect of chemoradiotherapy is to different trial designs and for completeness, the analyses were repeated including trials that used hydroxyurea or extended-field radiotherapy in the control arms. (Chemoradiotherapy for Cervical Cancer Meta-analysis Collaboration 2008) [1] |
| | | | *From example study 3: advanced bladder cancer*<br>In addition to the planned analyses described, we conducted supplementary analyses to investigate some |

© 2015 American Medical Association. All rights reserved.

| | | | |
|---|---|---|---|
| | | | of the previous criticisms of these trials in more detail. To assess whether modest imbalances impact on (a) the results of individual trials and (b) the pooled results over all trials, we performed Cox regression analyses, stratified by trial, including in the model terms for age, sex, grade, pT and pN categories. Because data on every variable were not available for all patients from each trial, a proportion of patients were necessarily lost from these analyses. Therefore, we conducted a second, unadjusted Cox regression analysis stratified by trial based on the same subset of patients, so that direct comparisons could be made. To investigate whether the collection and analysis of updated follow-up was able to counter any potential effects of early stopping in these trials, we estimated HRs from the trial publications using the reported statistics or from the survival curves [20] and compared these with HRs obtained from updated IPD. (Advanced Bladder Cancer Meta-analysis Collaboration 2005). [3] |
| **Results** | | | |
| Study selection and IPD obtained | 17 | Give numbers of studies screened, assessed for eligibility, and included in the systematic review with reasons for exclusions at each stage. Indicate the number of studies and participants for which IPD were sought and for which IPD were obtained. For those studies for which IPD were not available, give the numbers of studies and participants for which aggregate data were available. Report reasons for non-availability of IPD. Include a flow diagram. | *From example study 1: cervical cancer*<br><br>We identified 25 randomized trials that were eligible for the main analysis. We were unable to include data from 10 trials (1,113 patients), either because data could not be located (six trials, 814 patients) or because we were unable to make contact with the relevant investigators (four trials, 299 patients). Data were therefore available for 3,452 women from 15 trials. This includes 85% of women from trials that used cisplatin-based chemoradiotherapy and almost 80% of women from trials that used fluorouracil (FU)- and/or mitomycin-based chemoradiotherapy. (Chemoradiotherapy for Cervical Cancer Meta-analysis Collaboration 2008). [1] |
| | | | *From example study 4: radiotherapy in lung cancer*<br><br>The different steps of the trial selection are presented in {Figure 1}. Twelve eligible trials were identified, two in SCLC and 10 in NSCLC. Excluded trials are listed {Data Supplement}. Data were no longer available for two NSCLC trials so that 10 trials were analyzed, two SCLC trials and eight NSCLC trials. One trial had a factorial design: patients were also randomly assigned to receive or not concomitant CT; one trial had a randomization stratified on administration of induction chemotherapy. Each of these two trials was split into two separate trials, with and without chemotherapy. Therefore, 12 trials and 2,685 patients were analyzed {Table 1}. (Mauguen A. et al. 2012). [4] |
| | | | *Example study 6: spinal fusion*<br><br>The YODA project team provided IPD from 17 Medtronic trials. Eleven of these were RCTs comparing rhBMP-2 with ICBG surgery and were eligible for inclusion in our principal evaluation of effectiveness. Of the others, 4 were single-group trials of rhBMP-2 and 1 used a different comparator. We included these only in our supporting consideration of adverse events. We did not consider 1 trial that was stopped early after recruiting only 3 patients. {Figure 1} shows search results. In addition to the trials supplied by |

© 2015 American Medical Association. All rights reserved.

| | | | |
|---|---|---|---|
| | | | Medtronic, we identified 2 eligible randomized trials not conducted by Medtronic that compared rhBMP-2 with ICBG surgery. We requested IPD from the authors and obtained them from 1 study. Data from the other trial, which involved 40 patients having single level bilateral posterior lateral interbody fusion, were unavailable. (Simmonds et al. 2013). [6] |
| Study characteristics | 18 | For each study, present information on key study and participant characteristics (such as description of interventions, number of participants, demographic data, unavailability of outcomes, funding source and if applicable duration of follow-up). Provide (main) citations for each study. Where applicable, also report similar study characteristics for any studies not providing IPD. | *From example study 1: cervical cancer*<br><br>Patient characteristics for the 15 trials are listed in {Appendix TableA1 (online only)}. Data on age were provided for all trials, data on histology and stage were provided for 14 trials, data on performance status were provided for 12 trials, and data on grade were available for nine trials. Data on pelvic node involvement and iliac node involvement were available for six trials, with para-aortic node involvement available for nine trials. On the basis of the available data, women were mostly between 35 and 64 years of age, with good performance status. They had tumors that were largely of squamous cell histology (89%), stage IIb (36%), or stage III (36%), and moderately differentiated (35%). However, as there was generally no central pathology review, the precise definition of tumor grade may vary from trial to trial. Three trials excluded women with involved para-aortic nodes and para-aortic nodal status was either uninvolved (48%) or unknown (51%)for the vast majority of the women from the remaining trials. (Chemoradiotherapy for Cervical Cancer Meta-analysis Collaboration 2008). [1] |
| | | | Note that these data are usually presented in a table, or in an appendix if space is limited. |
| IPD integrity | A3 | Report any important issues identified in checking IPD or state that there were none. | *Example study 11: post operative radiotherapy in non small cell lung cancer*<br><br>The updated results of the Italian trial within the meta-analysis are less extreme than when the trial was originally published […] This is attributable both to extended follow-up and that data checking procedures identified anomalies in the original published dataset, which were subsequently rectified. (Burdett & Stewart 2005). [11] |

© 2015 American Medical Association. All rights reserved.

| | | | |
|---|---|---|---|
| Risk of bias within studies | 19 | Present data on risk of bias assessments. If applicable, describe whether data checking led to the up-weighting or down-weighting of these assessments. Consider how any potential bias affects the robustness of meta-analysis conclusions. | *Example study 6: spinal fusion*<br><br>Our assessment of risk of bias was the same for all Medtronic trials. Randomization and allocation concealment procedures were adequate for all trials. Neither patients nor physicians were blinded to the treatment received, and all pain and function outcomes were patient-assessed, so there was a potential for bias in these outcomes. Successful fusion was assessed by researchers blinded to the treatment received. (Simmonds et al. 2013). [6] |
| | | | From example study 2: preoperative chemotherapy in non small cell lung cancer [30]<br><br>Any risk of bias associated with the randomization procedure and completeness of outcome data in these 15 trials was judged to be low and the effects of early stopping were minimized by the collection of updated follow-up and investigated in the analyses. (NSCLC Meta-analysis Collaborative Group 2014). [2] |
| Results of individual studies | 20 | For each comparison and for each main outcome (benefit or harm), for each individual study report the number of eligible participants for which data were obtained and show simple summary data for each intervention group (including, where applicable, the number of events), effect estimates and confidence intervals. These may be tabulated or included on a forest plot. | Results of individual studies are commonly presented in a forest plot and described in the context of the overall synthesis. |
| Results of syntheses | 21 | Present summary effects for each meta-analysis undertaken, including confidence intervals and measures of statistical heterogeneity. State whether the analysis was pre-specified, report the numbers of studies and participants and, where applicable, the number of events on | *From example study 16: advanced ovarian cancer*<br><br>For comparison I, data were available from 16 of 22 eligible trials, comprising 1379 patients randomized to receive a single non-platinum agent and 1767 to receive non-platinum combinations. The reason for this imbalance in numbers was that some trials used two or more combination arms. A total of 2817 deaths were observed and the median duration of follow up was 10 years. Figures 1 and 2 show the results. There was no evidence of any overall difference between the two types of treatment (chi square = 0.65; p=0.42), the overall relative risk being 0.98 (95% confidence interval 0.91 to 1.05). (Advanced Ovarian |

© 2015 American Medical Association. All rights reserved.

| | | | |
|---|---|---|---|
| | | which it is based. | Cancer Trialists Group 1991). [15] |
| | | When exploring variation in effects due to patient or study characteristics, present summary interaction estimates for each characteristic examined, including confidence intervals and measures of statistical heterogeneity. State whether the analysis was pre-specified. State whether any interaction is consistent across trials.<br><br>Provide a description of the direction and size of effect in terms meaningful to those who would put findings into practice. | *From example study 2: preoperative chemotherapy in non small cell lung cancer*<br><br>We did not identify clear evidence that the effect of preoperative chemotherapy on survival differed by age, age group, performance status, or histology {figure 3}. Although, overall, there is no evidence of a difference in effect by sex, there is heterogeneity in the interaction {figure 3}. Some trials suggest the effect might be greater in women and others in men, but it is not clear why. Also, there was a significant interaction between the effect of preoperative chemotherapy and stage in the ChEST trial,32 but not in the other trials, or across all trials (interaction p=0·83; appendix). An exploratory analysis, splitting clinical stage I disease into IA and IB, also identified an interaction between the treatment effect and clinical stage in the ChEST trial, but not across trials (p=0·64, heterogeneity p=0·22). Thus, the overall HR of 0·87 was applied to the control group survival for each stage, giving an absolute survival improvement at 5 years of 5% for all stages, taking it from 50% to 55% in stage I, from 30% to 35% in stage II, and from 20% to 25% in stage III. However, most patients in stage I are IB (89%), in stage II are IIB (92%), and in stage III are IIIA (98%), therefore we can be most confident of results for these patients. (NSCLC Collaborative Group 2014). [2] |

© 2015 American Medical Association. All rights reserved.

| | | | |
|---|---|---|---|
| Risk of bias across studies | 22 | Present results of any assessment of risk of bias relating to the accumulated body of evidence, including any pertaining to the availability and representativeness of available studies, outcomes or other variables. | *From example study 3: advanced bladder cancer*<br><br>Overall, there was a slight imbalance by age across all trials, with a slightly higher proportion of younger patients in the chemotherapy arm than the in the control arm. This meant that when the model was adjusted for age alone, the estimate of effect moved towards equivalence compared with the unadjusted analysis. However, when all of the baseline characteristics (age, sex, grade, pT and pN) were taken into account, the Cox regression survival analysis tended more in favour of adjuvant chemotherapy, suggesting that overall, the proportion of poor prognosis patients was greater in the chemotherapy arm. (Advanced Bladder Cancer Meta-analysis Collaboration 2005). [3] |
| Additional analyses | 23 | Give results of any additional analyses (e.g. sensitivity analyses). If applicable, this should also include any analyses that incorporate aggregate data for studies that do not have IPD. If applicable, summarize the main meta-analysis results following the inclusion or exclusion of studies for which IPD were not available. | *From example study 2: preoperative chemotherapy in non small cell lung cancer*<br><br>Although this meta-analysis included most patients known to have been randomized, four eligible trials (198 patients) could not be included. We could estimate an HR for survival for one trial of 90 patients, but not the remaining three trials. Two of these (106 patients) did not report the appropriate information, and one (two patients) was unpublished. When the single estimated HR was combined with the overall result for the meta-analysis, the effect on survival remained the same (HR 0·87, p=0·006), but being based on 96% of patients who were randomized, it provides more convincing evidence of a benefit of preoperative chemotherapy. This systematic review and meta-analysis will be updated if further eligible trials are identified. (NSCLC Meta-analysis Collaborative Group 2014). [2] |
| | | | *Example study 8: High-dose chemotherapy with stem cell transplant in Non-Hodgkin Lymphoma*<br><br>The effect of the trial using extended-field radiotherapy on the control arm (HR, 0.50; 95% CI, 0.37 to 0.67; P = 0.000006) also differed from that in the main analysis (test for interaction, P=0.004), with an absolute survival benefit of 21% (from 50% to 71%) at 5 years. Excluding those studies where individual patient data were not available the HR as an IPD analysis for overall survival increases (HR 1.14; CI 0.98 to 1.34). A funnel plot analysis was performed to investigate for publication bias or other biases. Although graphical asymmetry could be observed for OS and EFS indicating that studies with negative findings might be underrepresented, related linear regression tests did not show significance (data not shown). (Greb et al 2009). [8] |
| **Discussion** | | | |

© 2015 American Medical Association. All rights reserved.

| | | | |
|---|---|---|---|
| Summary of evidence | 24 | Summarize the main findings including the strength of evidence for each main outcome. | *Example study 6: rhBMP-2 for spinal fusion*<br><br>Our principal analyses were based on data from 1408 individual participants in 11 eligible RCTs, including all trials sponsored by Medtronic (published and unpublished) and 1 additional trial. We found the randomization procedures to be adequate in all trials, but participants were not blinded to treatment. Although assessment of some outcomes, such as radiologic assessment of fusion, was blinded, patient-reported outcomes related to pain were not. Follow-up was reasonably complete up to our final analysis time point of 24 months. Although there is some potential for bias associated with patient-reported outcomes, in general, we consider the body of evidence for comparative effectiveness to be strong.<br><br>We found clear evidence that rhBMP-2 improves rates of fusion compared with ICBG; however, the Medtronic definitions of fusion that we used may have been stringent given that only 69% of ICBG recipients achieved fusion within 24 months, which is lower than would be expected generally. Inconsistency across trials was high, with large $I^2$ values at all time points.<br><br>We also found that rhBMP-2 improves back pain and quality of life compared with ICBG at between 6 and 24 months after surgery. However, these improvements in pain fall below previously described, clinically meaningful thresholds (estimated as between 4 and 17 percentage points for ODI score and ≥5.4 points for SF-36 PCS ) [….]<br><br>The IPD also indicate that rhBMP-2 may be associated with an increased risk for cancer, with nearly double the number of new cancer cases compared with ICBG recipients. The overall absolute risk for cancer is low in both groups, however, so whether this increased risk is genuine is uncertain. Adverse event data in the literature raise concerns that rhBMP-2 may increase the risk for heterotopic bone formation, osteolysis, radiculitis, and retrograde ejaculation. However, these findings should be interpreted cautiously because they are based on only published nonrandomized studies, most of which provided little information about the comparability of groups. (Simmonds et al. 2013). [6] |

© 2015 American Medical Association. All rights reserved.

| | | | |
|---|---|---|---|
| Strengths and limitations | 25 | Discuss any important strengths and limitations of the evidence including the benefits of access to IPD and any limitations arising from IPD that were not available. | *From example study 1: chemoradiotherapy in cervical cancer*<br>Although this meta-analysis provides the most comprehensive and up-to-date summary of the effects of chemoradiotherapy and is based on a large number of women from the large majority of the international trials, IPD from 10 trials were unavailable and might impact on these results. Nine of these trials, including 891 randomly assigned patients, would contribute to the main analysis. Although HR estimates based on the publications of three unavailable trials suggest that their inclusion would not change the results, and all of the unavailable data would only contribute 20% more data to the main analysis, it is possible that inclusion of IPD from these trials could modify our estimate of effect to some degree. (Chemoradiotherapy for Cervical Cancer Meta-analysis Collaboration 2008). [1] |
| Conclusions | 26 | Provide a general interpretation of the findings in the context of other evidence. | *From example study 5: Otitis media*<br>Acute otitis media is one of the most common childhood infections. Evidence from conventional systematic reviews suggested that antibiotics provide only marginal benefit. Most international guidelines therefore recommended selective use of antibiotics for acute otitis media, especially in children aged 2 years or older. In children younger than 2 years, no consensus had been reached. Some guidelines recommend antibiotics for all these children, whereas others advised antibiotics only for children under 2 years if they are severely affected or have persistent signs of disease or related comorbidity. An IPDMA showed that antibiotics were most beneficial in children younger than 2 years of age with bilateral acute otitis media, and in children with both acute otitis media and otorrhoea. Most international guidelines have been revised according to this finding. (Rovers et al. 2006). [5] |
| | | | *From example study 2: preoperative chemotherapy in non small cell lung cancer*<br>Further questions regarding which drugs to use, the duration of chemotherapy, and if the effect might be modified by predictive genetic biomarkers will need to be answered by new or ongoing trials. Nevertheless, these results provide the most complete evidence so far of the effects of preoperative chemotherapy, showing a significant improvement in overall survival, time to distant recurrence, and recurrence-free survival. (NSCLC Meta-analysis Collaborative Group 2014). [2] |
| Implications | A4 | Consider relevance to key groups | *From example study 2: preoperative chemotherapy in non small cell lung cancer* |

© 2015 American Medical Association. All rights reserved.

| | | | |
|---|---|---|---|
| | | (such as policy makers, service providers and service users). Consider implications for future research. | The results so far seem to suggest similar effects with either preoperative or postoperative chemotherapy, giving a choice of treatment options. Clinicians might consider that preoperative chemotherapy is preferable for poorer prognosis patients with larger, more advanced stage tumors, less able to tolerate chemotherapy after surgery, or in regions where surgery waiting lists are longer. Postoperative chemotherapy might be preferred by surgeons and by patients wishing to have potentially curative treatment immediately, or for those with earlier stage disease. It also allows for more reliable pathological staging to establish if subsequent chemotherapy is appropriate. (NSCLC Collaborative Group 2014). [2] |
| Funding | | | |
| Funding | 27 | Describe sources of funding and other support (such as supply of IPD), and the role in the systematic review of those providing such support. | *From example study 2: preoperative chemotherapy in non small cell lung cancer*<br><br>The sponsors of the study had no role in study design, data collection, data analysis, data interpretation, or writing of the report. […] The MRC Project Management Group was funded by the UK Medical Research Council and the IGR Project Management Group was supported by Institut Gustave-Roussy, Programme Hospitalier de Recherche Clinique (AOM 05 209), Ligue Nationale Contre le Cancer, and Sanofi-Aventis (unrestricted grants). (NSCLC Collaborative Group 2014). [2] |
| | | | *From example study 4: Radiotherapy in lung cancer*<br><br>Supported by unrestricted grants from French Programme Hospitalier de Recherche Clinique, Ligue Nationale Contre le Cancer, and sanofi-aventis (J.-P.P.). The funding sources had no role in study design, data collection, data analysis, data interpretation, or manuscript writing. (Mauguen A. et al. 2012). [4] |

**eTable 2: Examples of reporting systematic reviews and meta-analysis of IPD**

[a]Reproduced with permission of the PRISMA IPD Group, which encourages sharing and reuse.

{} denotes references, figures, and tables as reported in the original cited text rather than material provided in this supplement.

© 2015 American Medical Association. All rights reserved.
